# Genome-wide survival study identifies a novel non-HLA donor-recipient genetic mismatch associated with kidney allograft survival

**DOI:** 10.1101/2025.11.07.25339560

**Authors:** Vincent Mauduit, Axelle Durand, Martin Morin, Kane E. Collins, Matej Roder, Peter J. van der Most, Killian Maudet, Nayane SB Silva, Olivia Rousseau, Ashwini Shanmugam, Edmund Gilbert, Graham Lord, Gianpiero L Cavelleri, Harold Snieder, Stephan J.L. Bakker, Pierre-Antoine Gourraud, Mathieu Ribatet, Géraldine Jean, Clarisse Kerleau, Magali Giral, Nicolas Vince, Martin H. de Borst, Ondrej Viklicky, Peter J Conlon, Sophie Limou

## Abstract

**BACKGROUND:** Despite a sharp rise in kidney graft short-term survival rates and better donor-recipient *HLA* matching, mid- and long-term survival have not sufficiently improved over the past decades. Several studies suggest that non-*HLA* factors could be involved in kidney allograft injury, but no validated marker has yet been identified. Here, we aimed at finding genetic variations and mismatches associated with graft function and survival.

**METHODS:** Using genome-wide strategies, we tested the recipients’ and donors’ common variations (SNPs and CNVs), and the donor-recipient genetic mismatches for association with 1-year kidney graft function and with time-to-death-censored kidney graft failure in a monocentric European cohort of 1,482 complete donor-recipient pairs. We validated our findings through a meta-analysis in two independent European cohorts gathering a total of 1,842 additional complete pairs.

**RESULTS:** We did not identify any significant association with 1-year graft function. However, we discovered four non-*HLA* mismatches (3 SNPs and 1 CNV) associated with time-to-kidney graft failure. One signal in a regulatory region upstream the *TOM1L1* gene (p=6.3×10^-9^, HR=4.1) was successfully replicated in the validation cohorts (p*_meta_*_-analysis_=6.7×10^-9^, HR=2.9) and ranked among the top 50 rejection-specific genes in a pan-organ transcriptomics study. This locus was also associated with time-to-cellular and humoral rejection (p=0.02) in the discovery cohort in patients achieving primary graft function.

**CONCLUSIONS:** By running one of the largest ever performed kidney transplantation genomic analyses, we identified and confirmed a novel donor-recipient genetic mismatch in a biologically relevant non-*HLA* locus associated with kidney allograft failure.

**Lay Summary:** Despite significant advances in immunosuppression, kidney transplant recipients remain at risk of graft rejection and, in more severe cases, graft failure, which can lead to retransplantation, return to dialysis, or even patient death. Donor-recipient *HLA* compatibility is integrated into kidney transplant clinical care, as this genetic region is key for immunity and graft tolerance. However, several studies suggest that compatibility outside of the *HLA* region could also influence graft survival. We aimed at investigating this hypothesis in a cohort of 1,482 donor-recipient pairs and found a novel genetic region involved in kidney graft dysfunction that was validated after meta-analysis of two independent cohorts. These findings contribute to a better understanding of the impact of donor-recipient genetic compatibility on kidney transplant outcomes.

## Introduction

Kidney transplantation remains the best treatment for end-stage kidney disease, both in terms of survival and quality of life^1^. Despite major improvements in short-term graft survival (above 90% at 1-year in France^2^), mid- and long-term survival remains challenging (only 76% at 5 years for kidney transplants performed in France in 2017-2018^2^). Several donor and recipient factors are known to impact kidney transplant outcomes, such as age, medical history, comorbidities, and genetic factors with the importance of *HLA* (human leukocyte antigen) matching between donor and recipient. *HLA* genetic mismatches were previously repeatedly demonstrated to trigger immune responses targeting the graft, thus decreasing its survival^3,4^. However, those *HLA* mismatches fail to explain the whole phenotypic variability observed in kidney graft dysfunction^5^, and negative long-term outcomes can still occur even in the context of a perfect *HLA* matching^3^.

Over the past two decades, an increasing number of studies have thus investigated non-HLA antigens, or so-called minor histocompatibility antigens (mHA), involved in graft failure^6,7^. In a recent retrospective multicentric French cohort, *MICA* (MHC class I chain-related gene A) allelic mismatches and anti-MICA donor-specific antibodies (DSA) were associated with decreased graft survival^8^. Antibodies targeting mHA expressed by endothelial cells such as AT1R (angiotensin type 1 receptor)^9^ or ETAR (endothelin-1 type A receptor)^10^ have also been associated with increased allograft injury in several cohorts. The diversity of potential mHA targets on endothelial cells was recently demonstrated through a crossmatch assay in the context of early acute microvascular rejection^11^. Overall, these results underline that non-HLA genetic factors also play a role in kidney graft failure.

Genome-wide association study (GWAS) is a method testing for association between millions of genetic variants and a complex trait or disease. This approach has been implemented in kidney transplantation cohorts^12^ to identify new mHA targets by focusing only on the recipient’s genome^13,14^ or by jointly investigating the donor and recipient’s genomes^15,16^. Several post-transplantation outcomes, including kidney graft function, survival and acute rejection events^13–16^, have been assessed in the past few years. Overall, few non-HLA signals^13,14^ have been identified and the replication has proven challenging, including with the largest GWAS to date (n=2,094 pairs in a multi-centric design)^15^. These mixed results could be explained by the limited sample size from each center (from dozens up to 610 recipients), the relatively low number of total events (from 275 to 495), and the heterogeneity in time period (from 1981 to 2016), standard of care and phenotype definition.

The integration of biological knowledge in the genetic study design enabled several teams to emphasize the importance of non-*HLA* regions in donor-recipient compatibility. First, two groups focused on SNPs tagging for homozygous deletions in recipients who received an organ from a non-homozygous donor and identified a significant association with rejection for *LIMS1*^17^ and *CFHR*^18^ loci, respectively. Second, as SNPs identified by GWAS individually exhibit small to moderate effect sizes, a common strategy is to combine their effects into a polygenic risk score capturing the overall genetic burden. Based on the assumption that cell surface antigens play a role in anti-allograft immune responses, Mesnard *et al.* proposed to sum the number of amino-acid mismatches in transmembrane proteins from whole-exome sequencing data (n=53 pairs). They demonstrated a significant association of this score with the 3-year post-transplant kidney graft function, independently from *HLA* matching^19^. This non-HLA mismatch score was later adapted to GWAS data (n=477 pairs) by integrating non-synonymous SNPs within genes encoding transmembrane and secreted proteins, and significantly associated with graft failure^20^.

Here, we aimed at revisiting these findings by attempting to replicate the conclusions of the past decade in one of the largest homogeneous cohorts focusing on kidney transplantation genetics (n=1,969 pairs)^21^. We also aimed at investigating genome-wide donor-recipient incompatibilities for the first time by running SNP mismatch association studies. In our KiT-GENIE cohort, all transplants were performed in the same center in Nantes, France between 1999 and 2020, and backed by a rich and well curated database (DIVAT, *Données Informatisées and VAlidées en Transplantation*) containing precise clinical information, detailed follow-up and well-defined transplant outcomes. We thus conducted GWASs investigating the effects of SNPs and CNVs from European donors and recipients, and tested polygenic scores for association with two main kidney transplant outcomes: 1-year post-transplant kidney allograft function, and time-to-kidney allograft failure.

## Methods

### Study participants and phenotypes

All samples originally analyzed were part of the KiT-GENIE biobank^21^, which consists of donors’ and recipients’ genetic data and 200+ demographic and clinical variables from the DIVAT database^22^. All included kidney transplants were performed in the same center in Nantes, France between 1999 and 2020 (follow-up data available up to 2021). Patients and donors were eligible for enrollment in our study if consent for genetic testing and a DNA sample from both the recipient and the donor were available (see Supplementary Methods for detailed inclusion criteria). Two primary outcomes were studied: 1-year post-transplant kidney allograft function, which was calculated with the 2021 race-free CKD-EPI eGFR equation^23^ using creatinine measured during protocol follow-up visits and excluding patients with insufficient follow-up time; and time-to-death-censored kidney graft failure, which was defined as return to dialysis, preemptive retransplantation or death with a non-functioning graft. In addition, some signals of interest were also explored for rejection events that were defined as the first biopsy-confirmed antibody mediated or T-cell mediated rejection using the latest Banff classification on the day of biopsy review. Surveillance biopsies were performed at the 3-month and 1-year visits. We did not perform genome-wide analyses for 5-year eGFR and time-to-rejection due to a lack of statistical power for these outcomes in the KiT-GENIE cohort.

### Genotyping, quality control and imputation

Samples were genotyped using the Axiom PMRA chip (900K SNPs, Affymetrix). HLA typing was performed using standard protocols by local HLA labs and we had access to low-resolution (one-field) typing results. We performed the quality control steps, SNP, CNV and high-resolution *HLA* imputation according to the gold standard guidelines^24^ (see Supplementary Methods). We focused our analysis on individuals of European ancestry with residual genetic variance captured through principal component analysis (PCA) or a genetic relationship matrix (GRM) computation when using SAIGE. PCA plots of all included cohorts can be found in Figures S1-3. After imputation, a total of 8.2 million SNPs, 7,583 CNVs and 1,687 unique recipients, 1,642 unique donors and 1,482 complete donor-recipient pairs were available for subsequent analyses in the discovery KiT-GENIE cohort.

### Mismatch definition

Donor-recipient *HLA* epitopic mismatches were computed through HLA-EPI^25^ available on the Easy-HLA website, and were defined as the sum of *HLA-A*, *HLA-B*, *HLA-C*, *HLA-DRB1* and *HLA-DQB1* eplet mismatches. In addition, we defined non-*HLA* SNP allelic mismatches for complete donor-recipient pairs under a dominant model (0 or 1 code), *i.e.* when the donor exhibited one or two copies of an allele not found in the recipient’s genome (Figure S4) to assess the hypothesis of potential increased immunogenicity induced by the donor’s ‘new’ genetic material. This choice was also motivated by the fact that the latest combined non-HLA mismatch score from GWAS genotyping data used the very same dominant mismatch model^20^. For insertion/duplication CNVs, a mismatch was defined as the absence of the insertion/duplication in the recipient’s genome while present in the donor’s genome. Symmetrically for deletion CNVs, a mismatch was defined when the recipient was homozygous for the deletion while the donor was not (known as a “genomic collision model”). Here, we investigated both coding and non-coding mismatches in order to capture the possible immunogenic antigens, as well as the possible regulatory variants impacting the expression level of proteins important for alloimmune responses or graft injury.

Mismatches occurring in less than 1% of the pairs were discarded for the GWASs (268 variants with MAF>1% and a mismatch frequency <1%).

### Statistical analyses

Demographic tables were generated through the R gtsummary package (v.2.0.4). Chi-square and Fisher’s exact tests were applied for categorical variables and Wilcoxon rank sum tests for continuous variables. All SNP and CNV genome-wide analyses were performed at 3 different levels: recipients’ genetic data, donors’ genetic data, and combination of both through donor-recipient mismatches (see Figure S4). For 1-year post-transplant eGFR outcomes, GWASs were performed with multivariable linear mixed models implemented in SAIGE^26^. For time-to-graft failure, genome-wide survival studies (GWSSs) were performed using Cox proportional hazards models and the R Survival package (v3.2-11)^27^. We also tested the genomic collision model for CNV-tagging SNPs association with time-to-rejection as in the original paper^17^. Statistical significance was assessed with the Bonferroni threshold. For more details on model corrections and power analyses, see Supplementary Methods and Figures S5-S7.

### Polygenic scores computation

We computed the published chronic kidney disease (CKD) genome-wide polygenic score (GPS) constructed from kidney function GWASs^28^. We then fitted a logistic regression model using the R base stats package to test the association between the GPS and the CKD *vs.* non-CKD status as originally described. Additionally, we tested the linear association between the recipients’ and donors’ GPS with 1- and 5-year post-transplant eGFR, as well as their association with kidney graft survival using Cox proportional hazards models. We also investigated the previously published genome-wide non-*HLA* mismatch score that only focused on non-synonymous SNVs located in genes encoding secreted and transmembrane proteins^20^. We fitted a Cox proportional hazards model to test its association with time-to-graft failure as originally described. Details regarding the scores computation and models adjustments are available in the Supplementary Methods.

### Top signals validation

We had access to two independent European validation cohorts for assessing only the SNPs and CNV significantly associated with graft failure in the KiT-GENIE discovery cohort: (1) the United Kingdom and Ireland Renal Transplant Consortium (UKIRTC, n=1,477 pairs including 364 graft failure events) and (2) the Prague cohort from Czech Republic (n=365 pairs including 54 graft failure events. Similar quality controls, mismatch and phenotype definitions, and statistical models were used in each cohort as in KiT-GENIE (for more details, see Supplementary Methods and Table S1).

Each cohort’s summary statistics were pooled through a fixed effect meta-analysis using the R *metafor* package (v4.4)^29^.

## Results

### Description of the KiT-GENIE cohort

Table 1 gathers the main demographics and clinical characteristics for KiT-GENIE’s European recipients (N=1,674), donors (N=1,631) and full donor-recipient pairs (N=1,482) with available genetic data. The median post-transplantation follow-up time was 6.1 years in our cohort. Kidney allograft function data at 1-year were available for 92% of recipients (134 patients excluded due to insufficient follow-up time) with a median of 51 mL/min/1.73m^2^. For full pairs, there were 303 reported graft failure events (20%) with a median time of 5.1 years, and 171 (12%) transplants experienced at least one rejection event. Additional information on the KiT-GENE demographics can be found in the Supplementary Results and Table S2.

**Table 1.**
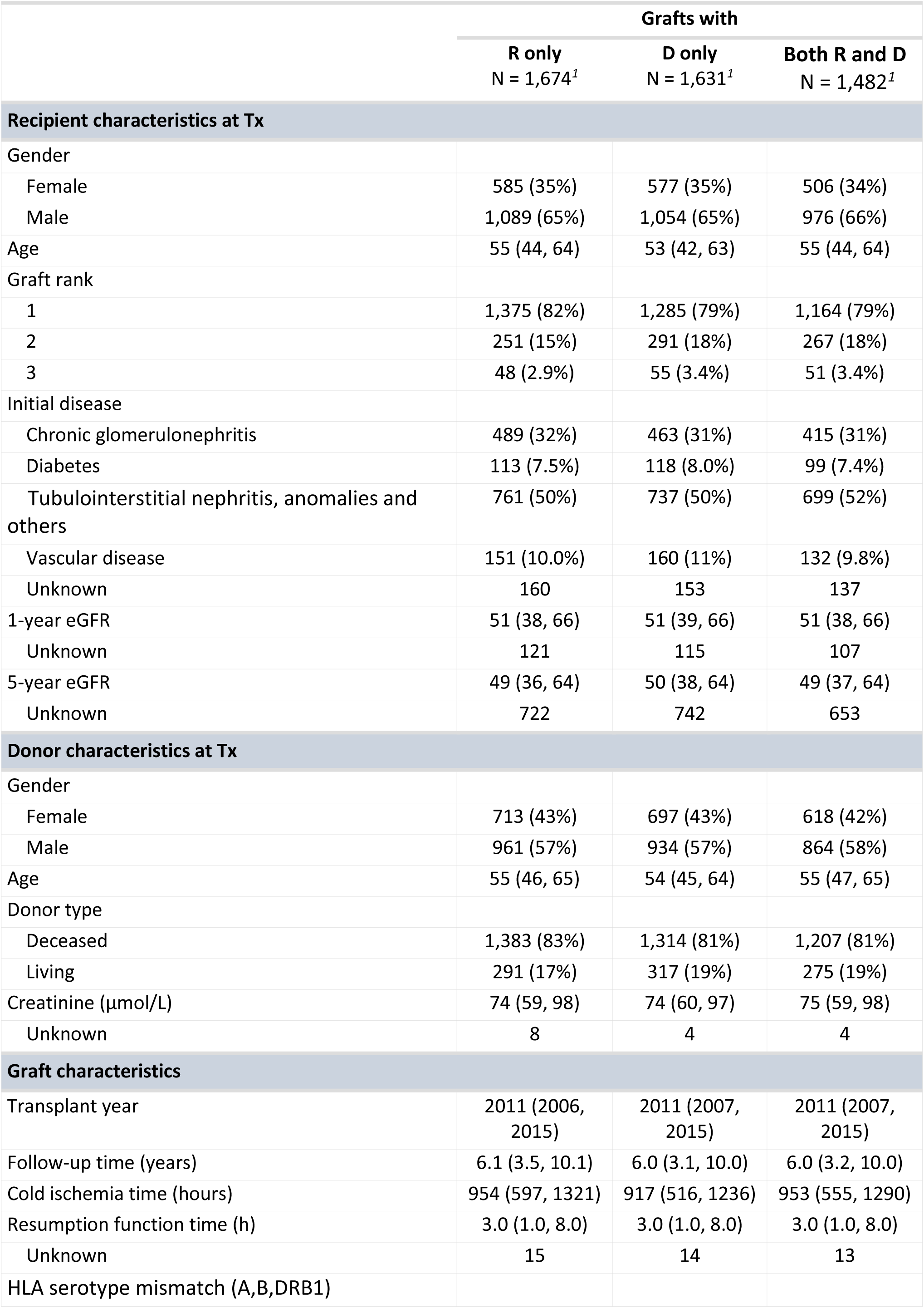
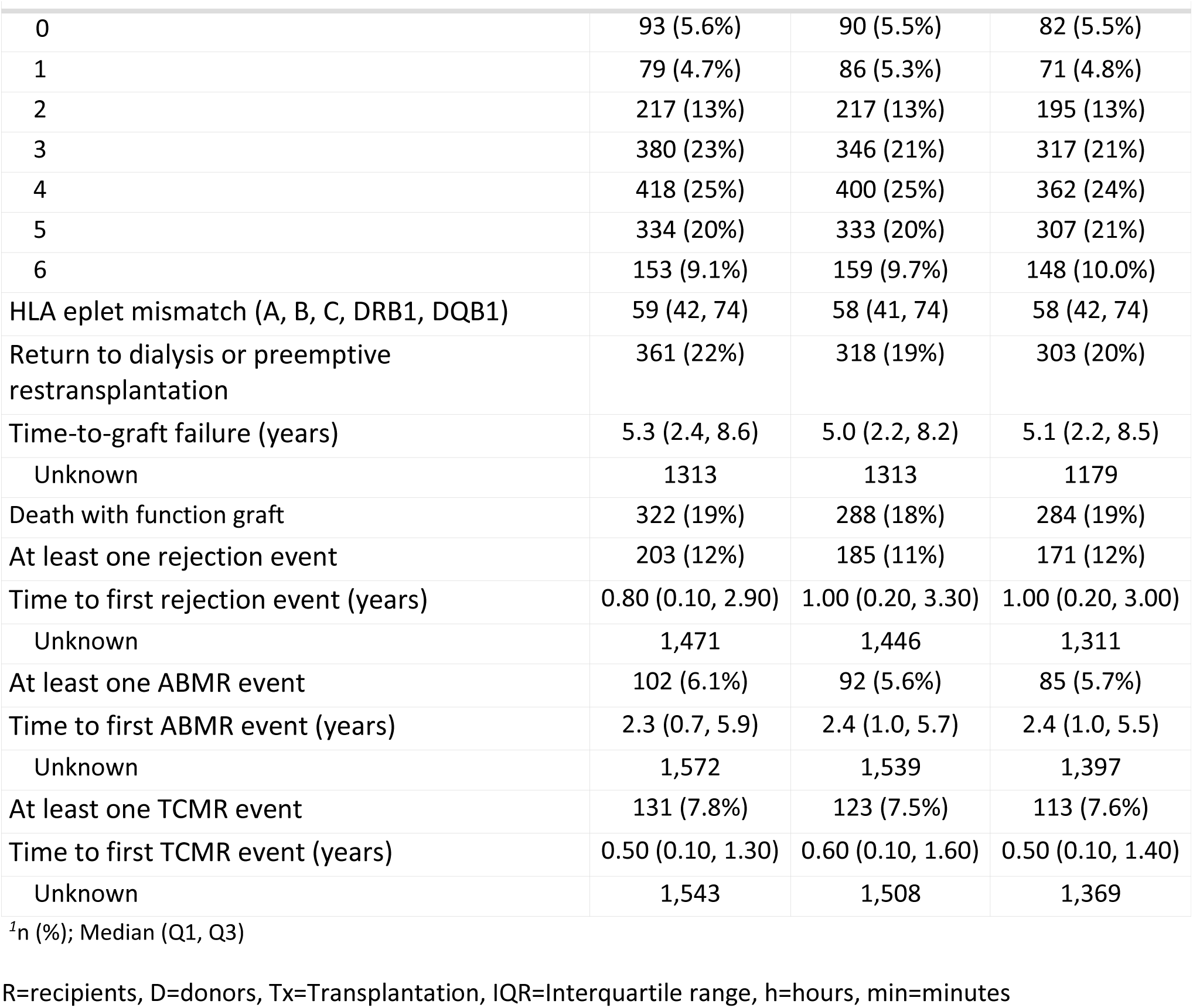
Demographic data for the KiT-GENIE cohort gathering recipients’, donors’ and graft characteristics.

|  | Grafts with |  |  |
| --- | --- | --- | --- |
|  | R only<br>N = 1,674<sup>1</sup> | D only<br>N = 1,631<sup>1</sup> | Both R and D<br>N = 1,482<sup>1</sup> |
| **Recipient characteristics at Tx** |  |  |  |
| Gender |  |  |  |
| Female | 585 (35%) | 577 (35%) | 506 (34%) |
| Male | 1,089 (65%) | 1,054 (65%) | 976 (66%) |
| Age | 55 (44, 64) | 53 (42, 63) | 55 (44, 64) |
| Graft rank |  |  |  |
| 1 | 1,375 (82%) | 1,285 (79%) | 1,164 (79%) |
| 2 | 251 (15%) | 291 (18%) | 267 (18%) |
| 3 | 48 (2.9%) | 55 (3.4%) | 51 (3.4%) |
| Initial disease |  |  |  |
| Chronic glomerulonephritis | 489 (32%) | 463 (31%) | 415 (31%) |
| Diabetes | 113 (7.5%) | 118 (8.0%) | 99 (7.4%) |
| Tubulointerstitial nephritis, anomalies and others | 761 (50%) | 737 (50%) | 699 (52%) |
| Vascular disease | 151 (10.0%) | 160 (11%) | 132 (9.8%) |
| Unknown | 160 | 153 | 137 |
| 1-year eGFR | 51 (38, 66) | 51 (39, 66) | 51 (38, 66) |
| Unknown | 121 | 115 | 107 |
| 5-year eGFR | 49 (36, 64) | 50 (38, 64) | 49 (37, 64) |
| Unknown | 722 | 742 | 653 |
| **Donor characteristics at Tx** |  |  |  |
| Gender |  |  |  |
| Female | 713 (43%) | 697 (43%) | 618 (42%) |
| Male | 961 (57%) | 934 (57%) | 864 (58%) |
| Age | 55 (46, 65) | 54 (45, 64) | 55 (47, 65) |
| Donor type |  |  |  |
| Deceased | 1,383 (83%) | 1,314 (81%) | 1,207 (81%) |
| Living | 291 (17%) | 317 (19%) | 275 (19%) |
| Creatinine (μmol/L) | 74 (59, 98) | 74 (60, 97) | 75 (59, 98) |
| Unknown | 8 | 4 | 4 |
| **Graft characteristics** |  |  |  |
| Transplant year | 2011 (2006, 2015) | 2011 (2007, 2015) | 2011 (2007, 2015) |
| Follow-up time (years) | 6.1 (3.5, 10.1) | 6.0 (3.1, 10.0) | 6.0 (3.2, 10.0) |
| Cold ischemia time (hours) | 954 (597, 1321) | 917 (516, 1236) | 953 (555, 1290) |
| Resumption function time (h) | 3.0 (1.0, 8.0) | 3.0 (1.0, 8.0) | 3.0 (1.0, 8.0) |
| Unknown | 15 | 14 | 13 |
| HLA serotype mismatch (A,B,DRB1) |  |  |  |
|  | R only<br>N = 1,674<sup>1</sup> | D only<br>N = 1,631<sup>1</sup> | Both R and D<br>N = 1,482<sup>1</sup> |
| 0 | 93 (5.6%) | 90 (5.5%) | 82 (5.5%) |
| 1 | 79 (4.7%) | 86 (5.3%) | 71 (4.8%) |
| 2 | 217 (13%) | 217 (13%) | 195 (13%) |
| 3 | 380 (23%) | 346 (21%) | 317 (21%) |
| 4 | 418 (25%) | 400 (25%) | 362 (24%) |
| 5 | 334 (20%) | 333 (20%) | 307 (21%) |
| 6 | 153 (9.1%) | 159 (9.7%) | 148 (10.0%) |
| HLA eplet mismatch (A, B, C, DRB1, DQB1) | 59 (42, 74) | 58 (41, 74) | 58 (42, 74) |
| Return to dialysis or preemptive retransplantation | 361 (22%) | 318 (19%) | 303 (20%) |
| Time-to-graft failure (years) | 5.3 (2.4, 8.6) | 5.0 (2.2, 8.2) | 5.1 (2.2, 8.5) |
| Unknown | 1313 | 1313 | 1179 |
| Death with function graft | 322 (19%) | 288 (18%) | 284 (19%) |
| At least one rejection event | 203 (12%) | 185 (11%) | 171 (12%) |
| Time to first rejection event (years) | 0.80 (0.10, 2.90) | 1.00 (0.20, 3.30) | 1.00 (0.20, 3.00) |
| Unknown | 1,471 | 1,446 | 1,311 |
| At least one ABMR event | 102 (6.1%) | 92 (5.6%) | 85 (5.7%) |
| Time to first ABMR event (years) | 2.3 (0.7, 5.9) | 2.4 (1.0, 5.7) | 2.4 (1.0, 5.5) |
| Unknown | 1,572 | 1,539 | 1,397 |
| At least one TCMR event | 131 (7.8%) | 123 (7.5%) | 113 (7.6%) |
| Time to first TCMR event (years) | 0.50 (0.10, 1.30) | 0.60 (0.10, 1.60) | 0.50 (0.10, 1.40) |
| Unknown | 1,543 | 1,508 | 1,369 |

<sup>1</sup>n (%); Median (Q1, Q3)
1 R=recipients, D=donors, Tx=Transplantation, IQR=Interquartile range, h=hours, min=minutes

### SNP association analysis

The time-to-failure GWSS was underpowered for finding low frequency variants (MAF<1%) with moderate effect size (Figure S6), and the 1-year eGFR GWAS was underpowered for identifying variants with a MAF<10% (Figure S7). No systemic inflation nor deflation was observed for the 1-year eGFR outcome on the QQplots (λ≤1.03), and only a minor inflation was detected for graft failure in the donor-recipient pairs analysis (λ=1.06, Figure S8). For kidney allograft function at 1-year post-transplantation, no genome-wide significant association was detected for the analyses focusing on recipients’ genome, donors’ genome or pairs’ genetic mismatches (Figures 1A-C). For the analyses on time-to-allograft failure, no significant association was detected with the recipient-restricted and the donor-restricted analyses (Figures 1D-E). Fourteen donor-recipient mismatches from three independent loci were significantly associated with time-to-graft failure (Figure 1F), in chromosomes 10 (rs75606962, p=2.0×10^-8^, HR=3.5 [2.3-5.4], mismatch frequency=3.2%), 17 (top rs16955257, p=6.3×10^-9^, HR=4.1[2.6-6.5], mismatch frequency=2.4%) and 21 (rs117176234, p=3.6.0×10^-8^, HR=5.1 [2.8-9.0], mismatch frequency=1.7%). The kidney graft survival was consistently significantly worse for the mismatched pairs than the matched pairs for these three loci in KiT-GENIE (Figures 2A and S9) according to the log-rank test (p<0.0001). Locus Zoom regional plots for chromosomes 10 and 17 signals are displayed in Figures S10-S11 to emphasize the linkage disequilibrium (LD) pattern around the top hits, whereas chromosome 21 signal was a singleton.

**Figure 1.**
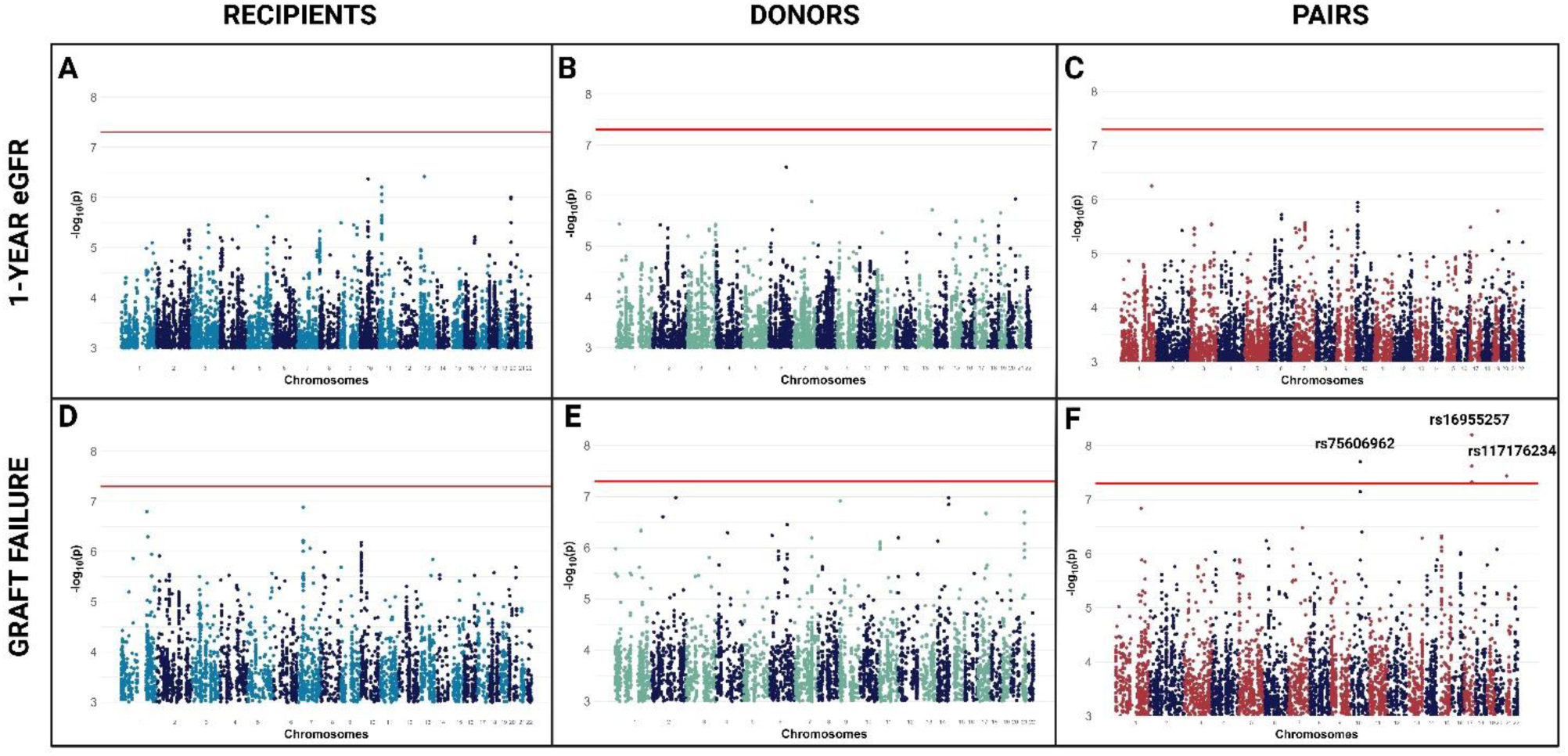
Summary of SNP association tests. Results of the association tests between recipients’ SNPs (panels A, D), donors’ (panels B, E) SNPs, and donor-recipient mismatches (panels C, F) with 1-year eGFR (panels A-C) and time-to-graft failure (panels D-F) in the KiT-GENIE cohort. The x-axis represents the variants’ position in the genome and the y-axis corresponds to the log-transformed p-values of the association tests. The Bonferroni genome-wide significance threshold is shown as a solid red line. Regression models were corrected for graft year, graft rank, recipient’s and donor’s age and sex, donor’s type (living or deceased), *HLA* epitopic mismatches and PCA-computed genetic ancestry. Three significant associations were identified for mismatches and time-to-graft failure (panel F) in chromosomes 10 (p=2.0×10^-8^, HR=3.5 [2.3–5.4]), 17 (p=6.3×10^-9^, HR=4.1 [2.5–6.5]) and 21 (p=3.6×10^-8^, HR=5.1 [2.9–9.0]). The chromosome 17 signal is composed of several significant SNPs in LD (r^2^>0.4): rs76528364, rs76466707, rs76132171, rs78872800, **rs16955257**, **rs78430798**, **rs80106929**, **rs79467893**, **rs78383882**, **rs117887990**, **rs77785449**, and rs7211161 (top SNPs in bold, these are in full LD).

**Figure 2.**
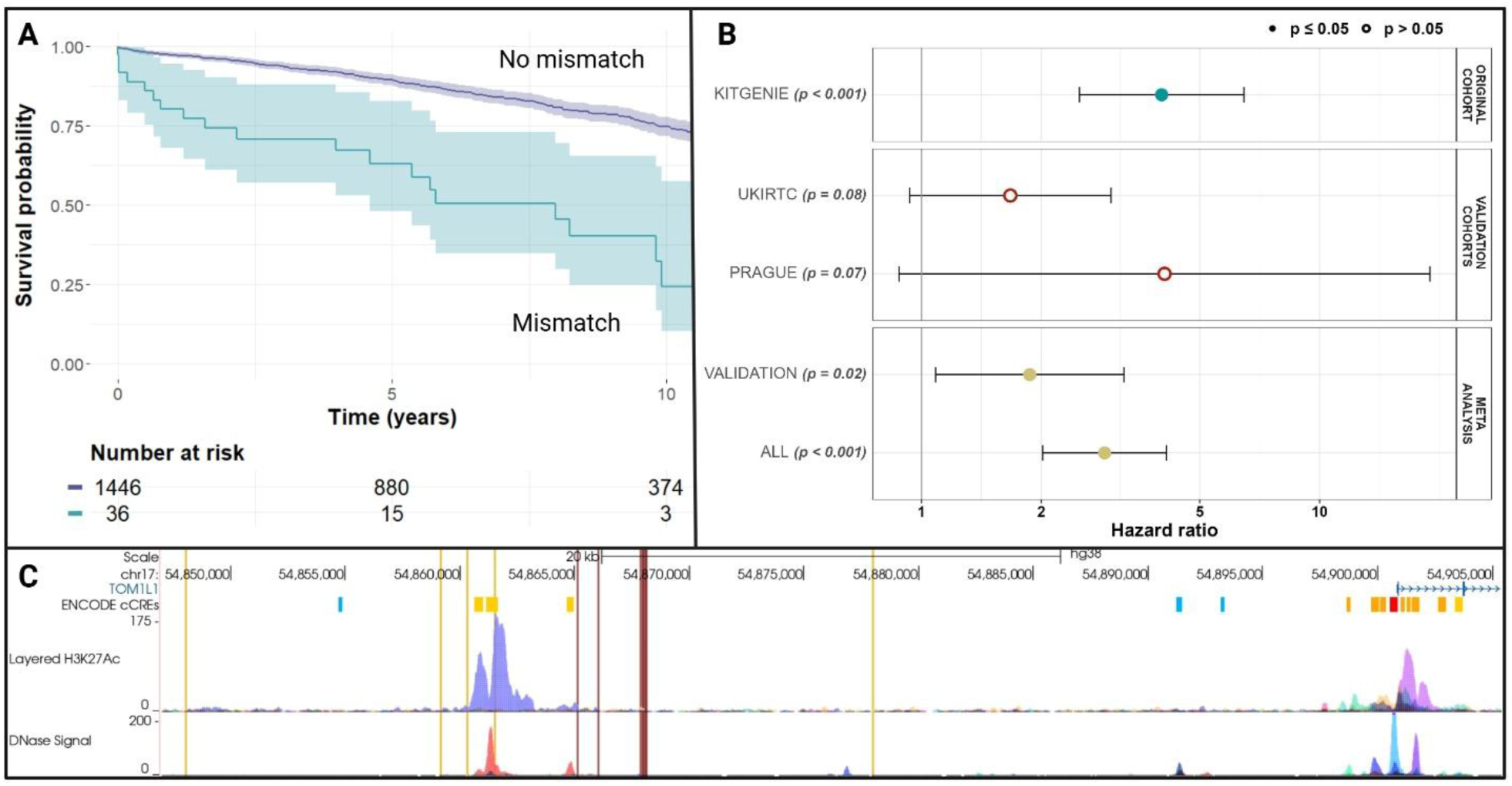
A replicated signal in chromosome 17. The top association between donor-recipient mismatches and time-to-kidney graft failure in the KiT-GENIE discovery cohort was located in chromosome 17. (A) Kaplan Meier plot comparing graft survival for the top signal (chr17, rs16955257) in KiT-GENIE. Mismatched donor-recipient pairs displayed a significantly worse graft survival than the matched pairs. (B) Forest plot showing hazard ratios (with 95% confidence interval, log-scale) of the mismatch in different cohorts. The association was tested in two independent validation cohorts. The signal did not reach nominal significance in individual validation cohorts (red dots). However, a combination of the two validation cohorts (yellow dot) through a meta-analysis yielded significance (p=0.02, HR=1.9 [1.4-3.2]). (C) Focus on the known regulatory elements within the top chromosome 17 signal region. Vertical red lines correspond to the top significant SNPs (7 SNPs in full LD) and vertical orange lines to the other significant SNPs, in LD with the top SNPs (r^2^>0.4). The signal overlaps with an open and active chromatin site (peaks in the layered H3K27Ac and DNAse tracks), and with several enhancers (marked in yellow on the ENCODE cCREs track) upstream the *TOM1L1* gene, which encodes a trafficking protein highly expressed in kidney and is part of a pan-organ rejection signature.

### Independent replication of the SNP mismatch findings

We tested these three SNP mismatch signals for replication in two independent validation cohorts. The variants’ frequencies were comparable between the three European cohorts (Tables S3-S5). Only the rs16955257 mismatch on chromosome 17 reached nominal significance in the validation meta-analysis (n=1,842 pairs) with an effect size in the same direction as in the KiT-GENIE cohort (p=0.02, HR=1.9 [1.4-3.2], Figure 2B and Table S6). The global meta-analysis of the discovery and validation cohorts exhibited strong statistical significance (n=3,324 pairs, p=6.7×10^-9^, HR=2.9 [2.0-4.1]). The leading rs16955257 variant is located 35kb upstream the *TOM1L1* gene (Figures 2C and S11).

### Lead chromosome 17 signal robustness and exploration

To identify potential genetic or non-genetic biases driving the lead variant association, we tested several iterations of the original Cox model and systematically found a hazard ratio of the same magnitude as in the original model, underlining the signal robustness (Table S7 and Supplementary Results). Notably, the association was driven by pairs where the donor brings the A alternative allele to the recipient, indicating that the donor’s brought genetic material may increase immunogenicity (p=1.3×10^-4^, HR=2.2 [1.5-3.4] when disregarding directionality and testing the donor-recipient genotype absolute difference, Table S7).

As graft failure is a heterogeneous outcome, we compared the causes of graft failure between the matched and mismatched groups and did not find a significant difference (p=0.05, Table S8). Chronic graft dysfunction was the first cause of graft loss in both groups. Graft failure events caused by vascular origin (21% vs 3.9%), disease recurrence (11% vs 6.3%) and rejection under immunosuppression treatment (11% vs 4.9%) were more prevalent in the mismatched group. To further assess the potential influence of non-genetic effects, we compared the matched and mismatched pairs in the discovery cohort for over 70 demographic and clinical factors (Table S9). No statistically significant difference was observed between the matched and mismatched group, although the mismatched group received nominally less tacrolimus at induction (72% vs 90%, p=1.9×10^-3^) and more MMF (86% vs 68%, p=1.9×10^-2^). However, the tacrolimus circulating levels were similar between the matched and mismatched groups over the follow-up course (Figure S12), indicating that the strength and adherence to the immunosuppression was similar in both groups. The mismatch association remained robust after adjusting for the induction treatment (tacrolimus and MMF, HR=4.0 [2.5-6.4], Table S7).

Although we did not find an association between the *TOM1L1* replicated mismatch and time-to-rejection (Table S7, p=0.62), we observed a nominally significant association for time-to-ABMR (58 events, p=0.02, HR=4.1 [1.3-13.8]) and time-to-TCMR (63 events, p=0.02, HR=4.2 [1.3-13.8]) in patients achieving primary graft function, suggesting this mismatch could be relevant for allograft outcomes beyond graft failure.

Finally, we ran *in silico* explorations on this replicated chromosome 17 signal to gain more insight on its biological relevance (Supplementary Methods). The frequency for rs16955257 and the SNPs in LD lies below the inclusion threshold for eQTL computations in most public datasets (MAF≥5%)^30^, so we currently lack data to directly characterize their functional impact on *TOM1L1* expression. However, we identified several kidney tissue-specific *TOM1L1* eQTLs close to the lead signal (Figure S11), suggesting that our chr17 variants could impact *TOM1L1* expression. In addition, several of the lead SNPs lay within an open and active chromatin site (peaks in the layered H3K27Ac signal and DNAse tracks, Figure 2C) with several enhancers (ENCODE cCREs track), which strongly indicates that our mismatch would capture a regulatory role. Lastly, the high conservation of the rs16955257-C allele across evolution (Figure S13) and the predicted pathogenicity of the A allele (CADD - Combined Annotation Dependent Depletion - score of 18.47, *i.e.* in the top 1.4% of all the human genome variants’ CADD score distribution) further suggests that this signal could have a functional pathogenic role. Additionally, querying 25 public transcriptomic datasets^31^ revealed that the TOM1L1 gene expression was systematically decreased in kidney graft during rejection.

### CNV association analysis

No systemic inflation nor deflation was detected (Figure S14). We reported a chromosome 2 duplication mismatch (HGSV_38325, tagged by the rs2392713 SNP, r^2^=0.87 in KiT-GENIE) significantly associated with time-to-kidney graft failure (Figures S15-16, p=1.1.x10^-6^, HR=4.5 [2.5-8.4], mismatch frequency=1.4%, which was not validated in the independent cohorts (p=0.70, Table S6, Supplementary Results).

In parallel, we assessed 39 deletion-tagging mismatches for association with time-to-rejection as originally tested^17,18^. Only the *LIMS1* rs893403 mismatch reached the multiple testing significance threshold in our cohort (p=0.001, HR=1.8 [1.3-2.6], Figure S17, Table S11, Supplementary Results).

### Genome-wide polygenic scores

First, the CKD GPS was not associated with CKD status (where the recipients represented CKD cases, p=0.7, Table S12) and the score distribution was similar between recipients and unrelated donors in our cohort (Figure S18). However, we found a nominally significant association between the recipient’s CKD GPS and 1-year post-transplant eGFR (p=0.04, OR=2.7 [1.1-6.7], Table S13, Supplementary Results).

Second, we computed the genome-wide non-*HLA* mismatch score as previously described^20^ (Figure S19) but did not replicate the association between the non-*HLA* mismatch score and time-to-kidney graft failure (p=0.14, HR=0.93 [0.84-1.03], Figure 3A and Table S13) despite having 3-times more pairs (n=1,482 *vs.* 477 pairs (39,420 SNVs *vs.* 59,268 in the original paper) and 100% statistical power (Table S14). We further implemented various iterations of the model, but the observed effect’s direction was almost systematically opposite to what was expected (HR<1, Table S15, Supplementary Results).

**Figure 3.**
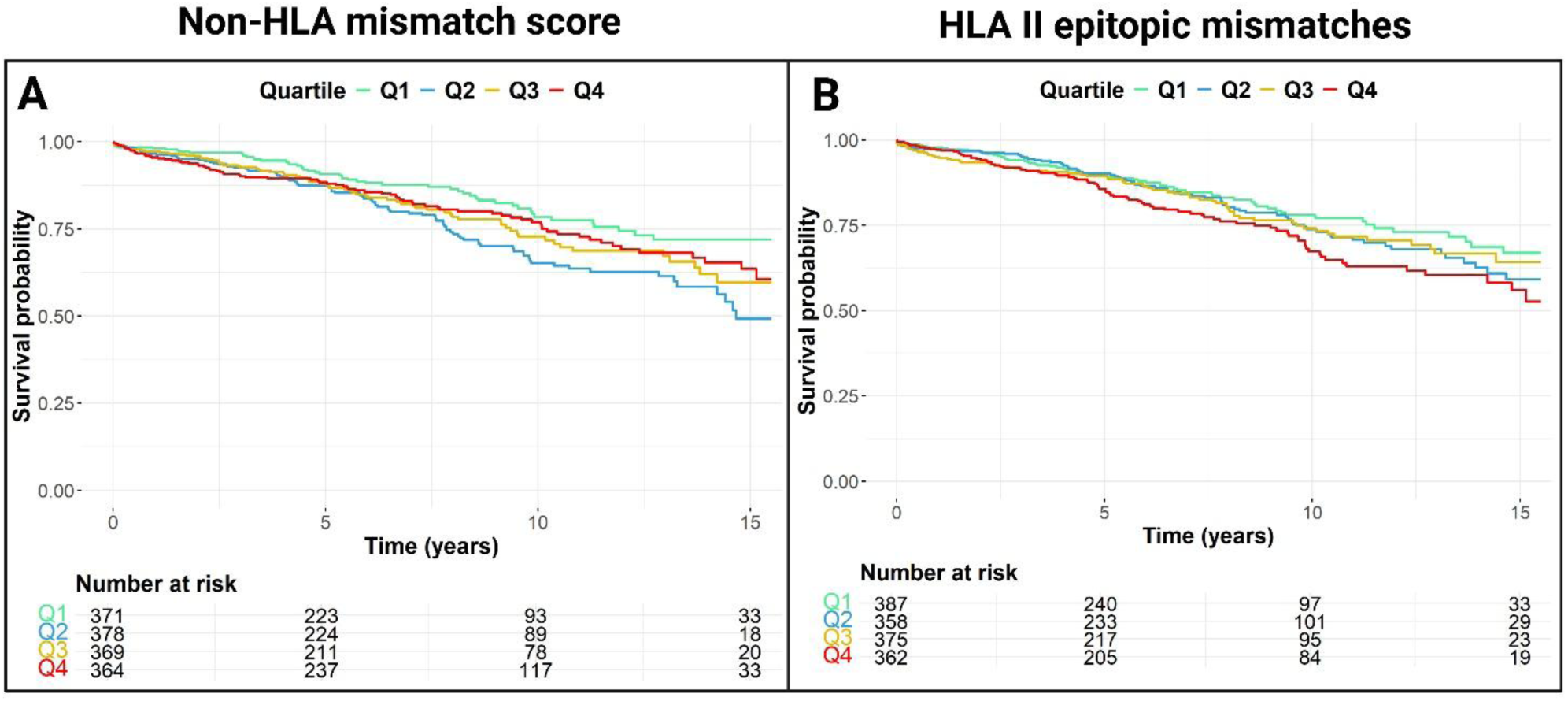
Survival curves for non-HLA and class II HLA mismatch scores. Kidney graft survival curves stratified by Reindl *et al.* non-HLA mismatch score quartiles (panel A) and by *HLA* II epitopic mismatches quartiles (panel B) in the KiT-GENIE cohort. While the highest *HLA* II quartile presents the worse survival (p=0.01), we did not identify a similar effect for the non-HLA mismatch score (p=0.14).

Finally, there was a nominal association between time-to-kidney graft failure and class II *HLA* epitopic mismatches (p=0.01, HR=1.16 [1.03-1.30], Figure 3B and Table S13), as well as for *HLA* ABDR mismatches (p=0.01, HR=1.10 [1.03,1.19]), but not with class I *HLA* epitopic mismatches (p=0.23, HR=1.08 [0.96-1.21]).

## Discussion

In this report, we aimed at investigating the recent findings of genetic studies revolving around kidney graft dysfunction and at conducting the first genome-wide investigation of non-HLA mismatches in a single large and homogeneous monocentric cohort (KiT-GENIE) with two independent validation cohorts (n_total_=3,324 European donor-recipient pairs), hence representing one of the largest studies ever performed in kidney transplantation genomics.

Firstly, our GWAS efforts did not replicate previous candidates or identify any individual SNPs or CNVs from the recipients or from the donors that were strongly associated with kidney graft function or time-to-graft failure, which supports the conclusions from a meta-analysis effort in 2,094 pairs^15^.

Secondly, we systematically investigated genome-wide donor-recipient mismatches at the SNP and CNV levels to assess the hypothesis of potential increased immunogenicity induced by ‘new’ genetic material brought by the donor. We investigated both coding and non-coding mismatches to capture the possible immunogenic antigens, as well as the possible regulatory variants impacting the expression level of proteins important for alloimmune responses or graft injury. CNV-tagging SNPs had previously been studied and two deletion-tagging mismatches were highlighted by two different groups in the *LIMS1*^17^ and *CFHR*^18^ loci. With a larger number of European pairs (n=1,482 *vs.* 705 and 1,025 pairs, respectively), we validated the *LIMS1* association with rejection, but not the *CFHR* one. Notably, another *LIMS1* haplotype mismatch was recently associated with kidney graft failure, further highlighting the potential importance of this genomic region as a mHA^32^. We also identified four novel donor-recipient mismatches (1 CNV mismatch and 3 SNP mismatch loci) significantly associated with time-to-kidney graft failure in chromosomes 2, 10, 17 and 21. Importantly, the three SNP mismatches lie within potential regulatory regions (*i.e.* introns or upstream genes) and in genetic loci that were previously among the most associated genes with CKD or a CKD-related trait: eGFR for chromosome 10 signal (p=2.4×10^-7^), gout for chromosome 17 (p=9.0×10^-21^) and CKD for chromosome 21 (p=4.7×10^-9^)^33^. This result underlines the importance of non-coding mismatches for kidney graft outcomes and the biological relevance of the identified loci.

### Biological and clinical relevance of the non-HLA mismatch findings

The chromosome 2 locus corresponds to a CNV duplication of 92 kb located between a block of genes and *EPHA4*. This genetic region was previously associated with immune responses and urate levels^34–37^. We did not have access to CNV data in the validation cohorts, so further work might be needed to replicate this signal. The chromosome 10 rs75606962 SNP mismatch is located in an intron of the *PALD1* gene encoding a tyrosine phosphatase. The rs75606962 SNP genotypes were previously correlated to gene expression levels of *TYSND1* and *SLC29A3*. *TYSND1* was reported with a decreased gene expression in CKD kidneys compared to normal kidneys^38^, and *SLC29A3* mutated mice exhibit an altered hematopoiesis and abnormal kidneys morphology, which emphasize the relevance of this whole genetic locus^39^. The chromosome 21 rs117176234 mismatch lies within an intronic region of *CHODL*, which encodes a chondrolectin previously associated with T-cell maturation and hypertension^40,41^. However, chromosomes 10 and 21 signal were not replicated in the validation cohorts (p=0.4 and p=0.7 respectively).

Finally, we successfully replicated the association with graft failure for the chromosome 17 mismatch (lead variant rs16955257) in two independent cohorts (n=1,842 pairs) through meta-analysis. This mismatch is located upstream *TOM1L1*, encoding a membrane trafficking protein with a clathrin-binding activity that is highly expressed in kidney cortex (median 10.6 transcripts per million) and medulla (16.1/million - notably the highest expression across all GTEx tissues)^42^. TOM1L1 protein expression is also reported in tubules in the Human Protein Atlas data^43^. Importantly, *TOM1L1* was reported to have a two-times lower expression in acute rejection kidney graft biopsies compared to stable graft biopsies^44^ and was identified as one of the 50 rejection-specific genes in a recent pan-organ transcriptomics study^31^, with consistently lower *TOM1L1* expression during organ rejection and fibrosis. In KiT-GENIE patients achieving primary graft function, we found a nominal association between the *TOM1L1* mismatch and both time-to-ABMR (p=0.02) and time-to-TCMR (p=0.02), suggesting a potential link with kidney transplant rejection in addition from graft failure. These observations will have to be confirmed in a larger cohort, considering the limited rejection rate in our cohort (12.2%, 58 and 63 events for ABMR and TCMR, respectively) and the fact that these outcomes were not available in the validation cohorts. Interestingly, our *in silico* explorations revealed that our top chromosome 17 signal is located close to *TOM1L1* kidney tissue-specific eQTLs^30^ and encompasses an open and active chromatin site comprising several enhancers upstream *TOM1L1*, which strongly indicates that our signal would capture a regulatory role. Additionally, the high conservation of the rs16955257-C allele across evolution and the predicted pathogenicity of the A allele further suggest this mismatch could drive a deleterious functional impact. If it remains challenging to interpret how a non-coding genetic mismatch can drive immunogenicity and affect graft survival, one could hypothesize that the A allele (present in the donor but not the recipient) decreases the enhancer activity, leading to a lower *TOM1L1* expression in the graft, which is associated with organ injury and rejection^31^. This differential expression could also induce immune responses, in the likes of autoantigens reactivity^45,46^. Overall, there is promising supporting biological evidence for our *TOM1L1* mismatch association, but functional experimental assays will be necessary to validate this hypothesis and to further disentangle the causal locus and pathophysiological mechanism at stake. This replicated chromosome 17 *TOM1L1* signal should therefore be a priority candidate for further functional investigations.

### Polygenic burden associated with kidney graft injury

Third, we investigated polygenic risk scores to capture the overall genetic burden associated with kidney allograft function and failure. Indeed, individual SNPs from the recipients or donors might only exhibit small to moderate effect sizes that are challenging to identify through a GWAS without very large sample size cohorts. When computing the CKD genome-wide polygenic score built from the general population in our cohort of kidney transplanted patients, we could not discriminate recipients who are CKD cases from unrelated donors who represent controls from the general population. However, higher recipients’ CKD scores were associated with lower 1-year post-transplant eGFR, without a sustained effect at 5-years. While polygenic risk scores are often underpowered for binary classification, these findings highlight the CKD score’s potential utility in the post-transplant context.

Then, we implemented a non-*HLA* mismatch score to capture all the non-synonymous mismatches from secreted and transmembrane protein-coding genes that could represent potential immunogenic antigens as previously proposed^19,20^. The underlying hypothesis is that each donor-recipient pair might carry different mismatch combinations and that the risk of kidney graft dysfunction and failure increases with a higher mismatch load. Using three times more pairs than the original paper (n=1,482 *vs.* 477) and benefiting from 100% statistical power for discovery, we unfortunately did not replicate the association between the non-*HLA* mismatch score and time-to-kidney graft failure, but we confirmed the well-described role of class II *HLA* mismatches. We applied the same model and the exact same SNP selection strategy to build the score as presented in the original manuscript, but we did not have access to the original list of variants. It is worth noting that Markkinen *et al.*^18^ did not replicate the association with time-to-graft failure either, while they found an association between the score quartiles and time-to-acute rejection in an adjusted model (we did not, Table S15).

Several reasons could explain this lack of replication. Firstly, graft failure is a heterogeneous outcome that is often caused by non-immune mediated factors^47^, which increases the potential effect size variability between cohorts. Secondly, although both European cohorts had a similar median follow-up (6.1 years for KiT-GENIE vs 5.3 for the original paper), other factors, including differences in adjustments for *HLA* in the allocation algorithm, immunosuppression regimens or period of inclusion (wider in KiT-GENIE during 1999-2020 vs 2005-2015 in the original paper), could have all contributed to the absence of replication. Thirdly, there was a substantial higher number of SNVs included in the original paper (39,420 *vs.* 59,268 SNVs). Even though the mismatch count was comparable between the two studies (1752 [IQR 1712-1798] vs 1892 [IQR 1850–1936] in the original paper), it is possible that some key variants were not present in our data. Lastly, Mesnard and colleagues emphasized the crucial role of rare variants in the original allogenomics mismatch score computed from WES data^19^. Despite great improvements in rare variants’ imputation, genotyping data may simply not be optimal for capturing refined donor-recipient non-HLA compatibility. In this line, further assessing the kidney transplantation genetic cohorts with next-generation sequencing might be critical. More studies are therefore necessary to build a robust and comprehensive non-*HLA* genetic score that would capture the genetic factors and mismatches impacting the risk of kidney graft failure. Integrating biological knowledge on genetic variant impact (*e.g.* non-synonymous variants, but also regulatory variants as highlighted in our SNP/CNV mismatch analyses), protein localization (*e.g.* transmembrane, secreted), or gene/protein tissue expression (*e.g.* immune cells *vs.* kidney from transcriptomics and proteomics assays) and regulation (*e.g.* epigenomics) could help refining such a score and should be compared to unsupervised strategies.

### Limitations

Despite our efforts, our study presents some limitations and we can wonder why, overall, no more replicated signals have been identified in kidney transplant genetics studies so far. Firstly, sample size is key when conducting omics studies, and even though KiT-GENIE is large in comparison to the kidney transplantation standards, GWASs tend to require thousands of cases for detecting low or moderate effect sizes. Larger sample sizes are especially essential when the associated genetic variants present with low frequency, as for our 3 significant SNP mismatches. It is important to underline here that our study was underpowered for identifying low frequency variants with moderate effect size and that the exclusion of patients with graft failure from creatinine-based analyses may have introduced a survival bias. Capturing variants with low effect size on kidney graft injury will likely require a large meta-analysis effort from the community. Secondly, graft failure, the primary phenotype examined in this study, is characterized by considerable heterogeneity^47^. There is indeed a wide diversity of possible graft failure causes, both immune and non-immune related, making it a suboptimal outcome for new association discovery. While the replicated signal’s matched and mismatched groups were similar for a wide a range of non-genetic factors and the association remained robust after several additional adjustments (Table S7), some moderate differences were observed in terms of graft failure cause. However, the challenges in determining and interpreting graft failure causes, the fact that more than one cause can contribute to graft loss^47^, and the limited sample size within our top signal mismatched group (n=19 events) prevented us from further stratifying our analysis for graft failure cause. We initially considered using kidney transplant rejection as the primary outcome for this study, as it might seem less heterogeneous than graft failure, but we lacked statistical power for genome-wide association discovery (5% for HR=2 and MAF=2%) due to the low rejection rate observed in our cohort (12%)^48^. Thirdly, there are potential unaccounted for confounding factors that might restrict our study power. Even though we tried to systematically mitigate biases, the monocentric nature of the KiT-GENIE cohort may limit the generalizability of some of our results. Our study time period remains large (from 1999 to 2020), which inevitably induces some heterogeneity in terms of standard of care and patient management (*e.g.* immunosuppression standards, crossmatch assay). Lastly, the leading signal’s hazard ratio magnitude varies greatly between the discovery and one of the validation cohorts, notably for UKIRTC that has comparable sample size and genetic ancestry background. While the mismatch frequency (2.4% vs 1.7%), event rate (20% vs 25%) and proportion of living donor transplants (19% vs 27%) were somewhat comparable, UKIRTC’s transplants are significantly older than KiT-GENIE’s (median transplant year is 2011 in KiT-GENIE vs 2001 in UKIRTC), which inevitably induces differences in *HLA* compatibility adjustments in the allocation system, patient management and immunosuppression regimen. Unfortunately, granular clinical and phenotypic data were not available in the validation cohorts to directly test for other biases. Beyond the inter-cohort heterogeneity, the observed hazard ratio’s discrepancies could also be attributed to a phenomenon known as the winner’s curse, which corresponds to an overestimation of effects ascertained by thresholding^49^. In this line, a direct genome-wide meta-analysis of the kidney transplant cohorts, even though significantly more time-intensive, would provide better effect size estimates and improve the findings’ robustness.

Overall, our study confirmed the absence of strong associations for individual common SNPs from the recipients or from the donors with short-term kidney allograft function and with time-to-graft failure. We did not replicate the association for the *CFHR* CNV mismatch and for the non-HLA mismatch score with kidney transplantation outcomes. However, we confirmed the role of class II *HLA* mismatches, we replicated the *LIMS1* deletion association with time-to-rejection and thanks to the integration of donor-recipient genetic compatibility, we discovered 4 non-HLA mismatches associated with time-to-graft failure, including one chromosome 17 signal that was replicated in independent cohorts. As only the integration of donor-recipient matching data led to significant associations for low frequency non-coding variants, we believe our results advocate for further studies in the kidney transplantation genetics field through sample size increase, donor-recipient pairs analysis, more refined phenotype definitions (*e.g.* specific rejection subgroups or histologic lesions), biological knowledge integration and by implementing next-generation sequencing efforts. In this regard, KiT-GENIE is part of the global data aggregation and meta-analysis effort of the transplant genetics community within the iGeneTRAiN consortium^50^, whose main objectives are to run large meta-analyses of kidney transplantation and cross-organ transplantation outcomes (including graft survival and rejection). We expect these on-going perspectives will lead to a better delineation of molecular mechanisms underlying progressive kidney graft dysfunction and failure.

## Acknowledgements

We thank everyone who helped in the design, collection, genotyping experiments, data cleaning and analyses. We are especially indebted to all donors and recipients who participated in this study, to the clinical staff and physicians who care and manage the patients on a daily basis and helped recruiting the patients, and to research associates who participated in the data collection. KiT-GENIE clinical data were collected from the French DIVAT multicentric prospective cohort of kidney and/or pancreatic transplant recipients by focusing on the Nantes centre (www.divat.fr, n° CNIL 914184, ClinicalTrials.gov identifier: NCT02900040). The analysis and interpretation of these data are the responsibility of the authors. We would like to acknowledge the local blood bank and HLA typing lab (*Etablissement Français du Sang* [EFS] Nantes) for facilitating the access to DNA samples. We thank the GenoBiRD and Curie genomic platforms for technical support and GWAS genotyping. We are also grateful to the Bioinformatics Core Facility BiRD, member of Biogenouest and *Institut Français de Bioinformatique* (IFB) (ANR-11-INBS-0013) for the use of their resources and their technical support. We would like to thank the *Centre de Calcul Intensif des Pays de la Loire*. We would like to thank the DNA laboratory of the department of immunogenetics, IKEM for preparing the DNA samples and the Department of Electronic Information Resources and Data Science, IKEM for preparing the clinical data of the Prague cohort.

The authors would like to thank the members of the DIVAT consortium for their involvement in the study, the physicians who helped recruit patients, and all patients who participated in this study. We also thank the clinical research associates who participated in the data collection. The analysis and interpretation of these data are the responsibility of the authors.

## DIVAT (Données Informatisées et VAlidées en Transplantation) Consortium

**Nantes :** Gilles Blancho, Julien Branchereau, Diego Cantarovich, Agnès Chapelet, Jacques Dantal, Clément Deltombe, Lucile Figueres, Claire Garandeau, Magali Giral, Caroline Gourraud-Vercel, Eloise Grenon, Clarisse Kerleau, Thibault Letellier, Christophe Masset, Aurélie Meurette, Simon Ville, Christine Kandell, Karine Renaudin, Florent Delbos, Alexandre Walencik, Anne Devis.

The authors would like to thank the iGeneTRAiN consortium (PI: B Keating) for introducing us to P Conlon and MH de Borst, and for integrating our cohorts into the on-going global collaborative meta-analysis effort in genomics of organ transplantation. We are pleased to have become active members of this initiative.

## Author contributions

VM and SL led the design and implementation of the primary analysis. VM and AD were involved in phenotypic data organization and preparation. MM was involved in the SNP imputation and the CNV analysis, realized the CNV imputation as well as the KiT-GENIE and Prague genotyping data quality control. GL and PC established the UKIRTC consortium. KC, AS, EG, PC, GC, PVDM, MDB, HS, SB, MRo and OV were in charge of the replication cohorts and analysis. KM computed the non-HLA mismatch score. OR, GJ, MRi and PAG helped on the statistical analysis and figure generation. NSB and NV were involved in the HLA imputation. MG and CK granted access to the DIVAT database. VM and SL wrote this paper. SL supervised the analysis, revised and formalized the scientific content of the article and mentored VM. All authors have read and approved the final version of the manuscript.

## Ethical approval

Written consent was obtained from each participant for access to clinical data and participation to DNA analyses (DIVAT n° CNIL 914184). Patients and/or the public were not involved in the design, or conduct, or reporting, or dissemination plans of this research. The UKIRTC was approved (or granted exemption) by the appropriate institutional and/or national research ethics committee which was the Hammersmith and Queen Charlotte’s & Chelsea Research Ethics Committee REC No 08/H0707/1.

## Data sharing

The GWAS aggregated data analyzed during the current study will be available for scientific review from the corresponding author upon reasonable request from the GWAS Catalog (GCP001044). Regarding subjects’ personal data under the responsibility of the sponsor, those data will be available for scientific review under the conditions defined by the EU General Data Protection Regulation. A data transfer agreement shall be signed with the sponsor defining the scope of the data transfer, as required by the GDPR and including an obligation to use the data for the sole purpose of the scientific review and forbidding their disclosure to third parties.

