## Supplementary for "Genome-wide survival study identifies a novel non-HLA donor-recipient genetic mismatch associated with kidney allograft survival"

**Supplementary Table S6 –** GWSS summary statistics for the SNP and CNV mismatches significantly associated with time-to-graft failure in the original KiT-GENIE cohort, and in the meta-analysis across the three validation cohorts. The chr2 CNV data were not available in the validation cohorts, so replication was assessed with the rs2392713 tagSNP (LD of $r^2$=0.87 in KiT-GENIE). The replicated chr17 SNP mismatch is highlighted in bold. Chr, chromosome; HR, hazards-ratio; VAL, validation cohorts. .. 22

**Supplementary Table S7** – Assessment of the replicated *TOM1L1* signal (rs16955257) robustness across various models, adjustments and subgroups from the KiT-GENIE discovery cohort. The summary statistics were obtained in the discovery cohort exclusively. *The original model is a Cox proportional hazards model fitted for time-to-failure and adjusted for graft year, graft rank, recipient and donor age and sex, HLA D-R epitopic mismatches, donor and recipient first four PCs. *Tx, Transplantation. GF, Graft Failure. ABMR, Antibody mediated rejection. TCMR, T-cell mediated rejection* ........................................ 23

**Supplementary Table S8** – Comparison of the reported graft failure cause between the *TOM1L1* chr17 matched and mismatched groups in the KiT-GENIE discovery cohort. A nominally significant difference was observed between the matched and the mismatched group, with a higher rate of failure events caused by disease recurrence and vascular disease in the mismatched group. We want to call for cautious in interpreting these results considering the low numbers of events for each cause (1≤n≤6) in the chr17 mismatched group. ........................................ 25

**Supplementary Table S9** - Demographic table comparing the chromosome 17 matched and mismatched groups for the replicated lead *TOM1L1* SNP signal (rs16955257) in KiT-GENIE for several non-genetic factors. When accounting for multiple testing ($P<7.1x10^{-4}$), no significant difference was observed. *Tx, transplantation.* ........................................ 31

**Supplementary Table S10 –** Frequencies for rs2392713, tagSNP for the chromosome 2 CNV signal in the KiT-GENIE discovery cohort ........................................ 31

**Supplementary Table S11 –** Genomic collision model replication for CNV tagging SNPs in the KiT-GENIE cohort. Results of the association tests between the high-priority deletion tagging SNPs selected by *Steers et al*. and time-to-rejection (39 of the 44 SNPs were present in KiT-GENIE). Mismatches were computed in two different ways: A1 homozygous recipient with A1 non-homozygous donors (columns 5-7), and A2 homozygous recipient with A2 non-homozygous donors (columns 8-10). The *LIMS1* deletion tagging SNP (rs893403, in bold), reached the adjusted Bonferroni threshold (p=0.001, HR=1.8 [1.3-2.6]), while the *CFHR* deletion tagging SNP (rs7542235, p=0.7) did not reach nominal significance. ........ 33

**Supplementary Figure S12 –** Evolution of the Tacrolimus circulating levels between the chr17 rs16955257 matched (red) and mismatched (blue) groups over the follow-up (missing data for N=180 patients out of 1,482). For each post-transplant visit (at 3, 6, 12 months and then annually thereafter), each curve represents the median Tacrolimus circulating levels within a group. Overall, median Tacrolimus circulating levels was of 7.1 ng/mL in KiT-GENIE and there was no major difference in the IS follow-up or adherence between the matched and mismatched groups. Data beyond 10 years is not shown here as the mismatch group sample size was very low. ........................................................... 45

**Supplementary Figure S13** – Multiple sequence alignment for the rs16955257 (in red) flanking regions across 45 non-human species. Sequences were retrieved from the RefSeq database and alignement was performed with the Clustal-O algorithm. ......................................................................................... 46

**Supplementary Figure S14 –** CNV association tests QQplots in the KiT-GENIE discovery cohort. No inflation nor deflation was detected. ......................................................................................... 47

**Supplementary Figure S15** - Summary of CNV association tests - Results of the association tests between recipients' CNVs (panels A, D), donors' CNVs (panels B, E), and donor-recipient CNV mismatches (panels C, F) with 1-year eGFR (panels A-C), and time-to-graft failure (panels D-F) in the KiT-GENIE cohort. The x-axis represents the CNVs' position in the genome and the y-axis corresponds to the log-transformed p-values of the association tests. The Bonferroni genome-wide significance threshold is shown as a solid red line. Regression models were corrected for graft year, graft rank, recipient's and donor's age and sex, donor's type (living or deceased), *HLA* epitopic mismatches and PCA-computed genetic ancestry. A significant association was identified between a CNV mismatch in chromosome 2 and time-to-graft failure (panel F, $p=1.1x10^{-6}$, HR = 4.5 [2.5 – 8.4]). ............................ 48

**Supplementary Figure S16 -** A chromosome 2 duplication mismatch (CNV tagged by the rs2392713 SNP) was significantly associated with time-to-kidney graft failure in the KiT-GENIE discovery cohort (p=1.1x10-6, HR=4.5 [2.5-8.4]). (A) The Kaplan Meier plot displays that mismatched pairs have a worse graft survival than matched pairs. (B) UCSC genome browser view of the genetic region surrounding the duplication (solid red line). The insertion lies in an intergenic region 900 kb downstream the EPHA4 gene. ...................................................................................................................... 49

**Supplementary Figure S17 –** Survival curves stratified by the deletion-tagging SNP mismatches in the *LIMS1* locus (left panel) and the *CFHR* locus (right panel) for the KiT-GENIE cohort. The *LIMS1* mismatch confers a significant worse risk of rejection (p=0.001), while we did not find a significant effect of the *CFHR* locus mismatch on rejection (p=0.69)........................................................................ 50

**Supplementary Figure S18 –** Boxplot comparing the distribution of the CKD GPS developed by Khan *et al*. between recipients (considered as CKD cases) and unrelated donors (considered as controls) in the KiT-GENIE cohort. We did not observe any major difference between recipients and donors (p=0.7). 50

**Supplementary Figure S19** - Distribution of the non-HLA mismatch score (before normalization) in unrelated (blue, coefficient of variation = 4.3%) and related (red, coefficient of variation = 12.2%) donor-recipient pairs from the KiT-GENIE cohort. The mismatch score is significantly lower in related individuals, as expected (Pearson correlation coefficient of -0.98 with the donor-recipient identity-by-descent value). ........................................................................................................................ 51

### Supplementary Methods

**KiT-GENIE inclusion criteria |** A total of 84% (2,632) of DIVAT transplants performed between 1999 and 2020 were included and both recipient's and donor 's DNA samples were processed to genotyping. For subsequent analyses, transplants were excluded if the recipient or donor was not an adult (< 18 years old), there was an immediate failure event, the graft rank was 4 or more, or in the case of a multi-organ transplant, to exclusively focus on kidney transplantation (529 transplants excluded in total). Additionally, non-European samples were removed (384 in total). In the case of repeated transplants, only the first known graft was kept for subsequent recipients-only analysis. For dual donation, one of the two transplants was randomly discarded for subsequent donors-only analyses. For donor-recipient interactions analyses, both repeated transplants and dual donation were kept, as the donor-recipient pair uniqueness is not compromised in this situation. Incomplete donor-recipient pairs reflect missing recipient or donor DNA (408 pairs removed), quality control (QC) failure (DNA QC, technological QC, genotyping data QC, 326 pairs removed) or donor/recipient not meeting the inclusion criteria (272 pairs removed).

**KiT-GENIE GWAS quality controls** | Samples were genotyped using the Axiom PMRA chip (Precision Medicine Research Array) which targets more than 900K variants and is enriched with immunological and functional variants. The primary technological QC steps were performed with Axiom Analysis Suite to assess quality metrics on plates, individuals and genetic variants[1]. A standard GWAS QC was then applied to remove variants violating the Hardy-Weinberg equilibrium ($p<10^{-6}$) or with a high level of missingness (>2%). All individuals (donors and recipients) respected the 3% missingness threshold, the genetic sex check, and the relatedness check. Inter-recipient (*resp.* inter-donor) relatedness up to 2$^{nd}$ degree was assessed with the KING[2] software using proportion of genomes sharing identity-by-descent >0.25. Ancestry was determined with RFMIX[3] using the 1000 Genomes reference panel (GRCh38, 30X Phase 3[4]).

Non-European individuals (defined with a global European ancestry averaged across autosomal chromosomes <0.9) were excluded from this study as the KiT-GENIE biobank consists mainly of Europeans (4030 out of 4414 samples, 91.3%). Residual genetic variance was captured through principal component analysis (PCA), or through a genetic relationship matrix (GRM) computation when using SAIGE.

SNP genotype imputation was performed through the TOPMed Imputation Server[5] using the TOPMed reference panel r2[6]. CNVs were imputed from genotyping data using Beagle[7] and the 1000 Genomes reference panel (GRCh38, 30X Phase 3). High-resolution (2-fields) *HLA-A*, *HLA-B*, *HLA-C*, *HLA-DRB1* and *HLA-DQB1* genotypes were obtained from SNP genotypes by imputation using HIBAG[8]. Only common (minor allele frequency > 1%) genotyped and high-quality imputed SNPs ($R^2$>0.8), CNVs ($R^2$>0.8) and *HLA* alleles (post-probability >0.5) were kept for subsequent analyses.

In total, filtering and GWAS quality controls removed 5,687 variants and 2 samples due to missing data. Hardy-Weinberg and minor allele frequency (MAF) filtering removed 277,380 variants. Post-imputation QC removed 36M variants through the 1% MAF filter and 660,298 variants through the imputation quality filter (R2>0.8).

**Statistical analyses |** For 1-year post-transplant eGFR, the multivariable linear mixed models were adjusted for graft year, graft rank, donor's age and sex, donor's type (living or deceased), *HLA* epitopic mismatches and GRM-computed genetic ancestry. As recipient's age and sex were incorporated in the eGFR equation, these were not included in covariates. For time-to-graft failure, the Cox proportional hazards models were adjusted for graft year, graft rank, recipient's and donor's age and sex, donor's type (living or deceased), *HLA* epitopic mismatches and PCA-computed genetic ancestry (*i.e.* first four principal components of recipients, donors or both if pairs were analyzed). For the genomic collision model, Cox proportional hazards models with time-to-rejection were adjusted for recipient and donor age and sex, *HLA* incompatibilities, donor type (living or deceased), donor and recipient ancestry (first four principal components) as in the original paper[11]. Rejection events were right-censored for death, lost to follow-up and graft failure.

The proportional hazards assumption was checked using the R Survival package (v3.2-11)[12] with the cox.zph function to test the correlation between the Schoenfeld residuals and time, and a p-value significance cut-off of 0.05. For pairs only, the proportional hazards assumption was violated for donor age (Figure S5), which was corrected by the implementation of an interaction term with time (log-transformed since the Schoenfeld residuals were slowly decreasing over time). Survival curves were computed through the Kaplan-Meier estimator. Competing risk models were computed using the R casebase package (v.0.10.6)[13].

Power analyses for the GWASs were performed with the R survSNP package (v.0.26)[14] for time-to-event data (Figure S6) and with R genpwr package (v.1.0.4)[15] for 1-year eGFR (Figure S7). For recipients and donors separate analyses, an additive model was used for simulation whereas a dominant model was used for pairs. For non-genome wide survival analyses, power estimations were performed with the R powerSurvEpi package(v.0.1.3)[16]. Finally, the R pwr package (v.1.3-0)[17] was used for general linear models.

To assess significance, we used the Bonferroni correction threshold ($p<5x10^{-8}$ for genome-wide analyses, 0.05/N for single markers association testing, where N is the number of independent tests).

**Computation of polygenic scores |** For computing the CKD GPS, we used the public list of SNPs and weights[18] and filtered variants with bcftools (v.1.14)[19]. Their weighted sum was computed through R base and the GPS was normalized according to the original paper's methods. We calculated the GPS on recipients and unrelated donors separately, and considered the kidney transplanted recipients as CKD cases while unrelated donors were considered as non-CKD controls. Related donor-recipient pairs (up to $2^{nd}$ degree) were indeed excluded from each model linked to the CKD GPS to prevent correlations between donor-recipient relatedness and score value. Models were corrected for diabetes, donor and recipient age and sex and the first four principal components as originally described.

Regarding the computation of the genome-wide non-HLA score, we used the strategy described in the original paper with variant annotation through ANNOVAR[20] and the UniProt database[21] (2024-03 release, https://www.uniprot.org/). Again, a mismatch was defined as the donor carrying an allele not exhibited by the recipient. No frequency cutoff was applied here and all SNVs/SNPs were considered, according to the original paper's methods. All mismatches were then summed to define the non-*HLA* mismatch score and we fitted a Cox proportional hazards model to assess its association with time-to-graft failure, adjusting for donor age and sex, donor type, *HLA* epitopic mismatches. We defined alternate non-HLA mismatch scores to further assess the relevance of some parameters, including the location of the SNVs (coding or not) and the transplantation context (*e.g.* deceased donors only, first graft rank only).

**Validation cohorts** | We had access to two independent European validation cohorts for assessing the SNPs and CNV significantly associated with graft failure in KiT-GENIE. First, the United Kingdom and Ireland Renal Transplant Consortium (UKIRTC) gathered 1,477 complete donor-recipient pairs (including 364 graft failure events) collected across UK and Ireland. UKIRTC data were collected from an earlier era than KiT-GENIE (mean transplant year=2001) and samples were genotyped using the Illumina 670 Quad Custom chip. Second, the Prague cohort comprised 365 complete donor-recipient pairs (including 54 graft failure events) from Czech Republic, and samples were genotyped using the Axiom PMRA chip. In both cohorts, the same QC and imputation pipeline were applied as for KiT-GENIE (TOPMed reference panel r2 for UKITRC and r3 for the Prague cohort). The same mismatch definition and statistical models as in KiT-GENIE were implemented with correction for *HLA-A*, *HLA-B*, *HLA-DRB1* mismatches instead of epitopic mismatches. For more details on each validation cohort, see the demographic Table S1.

**Top signal *in silico* exploration |** Human kidney-specific eQTLs were identified thanks to a recently published public resource created by the Susztak lab[24]. Importantly, the inclusion threshold was restricted to SNPs with a MAF>5% for their eQTL computation.

The regional plot in Figure 2C was generated through the UCSC genome browser[25]. The significant SNPs of the chromosome 17 signal, in LD ($r^2$>0.4) and represented as red (top SNPs) and orange vertical lines on Figure 2C, are: rs76528364, rs76466707, rs76132171, rs78872800, **rs16955257, rs78430798, rs80106929, rs79467893, rs78383882, rs117887990, rs77785449**, and rs7211161 (top SNPs in bold). They were overlayed with the open and active chromatine sites and enhancers tracks, which rely on public data generated by the ENCODE project[26].

The pathogenicity of the lead variant was assessed through the CADD score[27]. The CADD (Combined Annotation Dependent Depletion) pathogenicity score combines several metrics on conservation across evolution and on coding and non-coding annotations. As the high CADD score (*i.e.* in the top 1.4% of all the human genome variants' distribution) for the chromosome 17 appeared to be mostly driven by conservation metrics, we performed a multiple sequence alignment for the lead rs16955257 flanking regions across 45 non-human species. All genomic sequences were retrieved from the RefSeq database and the alignment was performed with the Clustal-O algorithm[28].

### Supplementary Detailed Results

**Characteristics description of the KiT-GENIE cohort |** Overall, the recipients were mostly male (65%) with a median age of 55 years-old at transplantation, and a vast majority (82%) benefited from their first kidney transplant over the study period. The donors were 83% deceased donors and 57% male with a median age at donation of 55 years-old. Ninety-four percent of kidney allografts did not involve an *HLA* A/B/DRB1 allelic full-matching between donor and recipient. Transplants were included for a period ranging from 1999 to 2020. Among full donor-recipient pairs, 156 were related up to $2^{nd}$ degree. The median post-transplantation follow-up time was 6.1 years in our cohort and 8% of the recipients were lost to follow-up (Table S2). The first-year visit was performed on average after 363 days (sd=26 days). Among the 134 patients who did not have 1-year follow-up data, 53 experienced a graft failure event in the first year, 49 died with a functioning graft, 5 were loss to follow-up and 27 either did not have sufficient follow-up or available 1-year visit information. Kidney allograft function data at 1-year and 5-year post-transplant were available for 92% and 56% of recipients, respectively (134 and 735 patients excluded due to insufficient follow-up time) with a median of 51 mL/min/1.73$m^2$ and 49 mL/min/1.73$m^2$, respectively. A total of 364 (22%) graft failure events were observed for KiT-GENIE recipients over the study period.

Supplementary Table S2 further presents the characteristics of both the control group and the cases of kidney graft failure events. As expected, the patients in the failure group were transplanted earlier (2008 vs 2012 median graft year), experienced more frequently at least one rejection event (26% vs 8%) and experienced more recurrence of the initial disease (8.6% vs 2.7%). Notably, the failure group received MMF more frequently at the beginning of the follow-up (87% vs. 63%) and tacrolimus less often (83% vs 92%), which reflects an evolution of protocols in the Nantes center over the study period.

**Lead chromosome 17 signal robustness and exploration |** As the rs16955257 mismatched group displays attrition early in the follow-up (Figure 2A), we performed a sensitivity analysis in patients achieving primary graft function after transplantation. The association was robust in this subgroup (Table S7, n=950 with 159 graft failure events, $p=3.9 \times 10^{-5}$, HR=4.3 [2.1-8.5]) with a higher p-value (likely due to the lower sample size leading to higher standard errors) but a comparable hazard ratio. The association remained significant when adjusting for rejection or disease recurrence, or when considering death as a competing event (HR>3.8, Table S7). Additionally, the signal also remained robust in the discovery cohort when adding further genetic covariates such as donor-recipient relatedness ($p=6.3 \times 10^{-9}$, HR=4.1 [2.6-6.5]) and donor-recipient PC1xPC2 Euclidian distance ($1.0 \times 10^{-8}$, HR=4.0 [2.5-6.4]). Importantly, the simultaneous inclusion of all the potential genetic and non-genetic confounding variables did not change the strength of the association ($p=3.1 \times 10^{-8}$, HR=3.9 [2.4-6.2], Table S7). When testing the association between the donor-recipient genotypes absolute difference (disregarding directionality) and time-to graft failure, we found a weaker effect (Table S7, $p=1.3 \times 10^{-4}$, HR=2.2 [1.5-3.4]), indicating that the association is driven by pairs where the donor brings an alternative allele to the recipient according to our hypothesis. Considering the frequency of the variant, the mismatch association is only driven here by donors bringing the rs16955257-A allele to homozygous CC recipients.

**CNV association analysis |** No CNV from the recipients, donors, nor pairs was significantly associated with 1-year post-transplant eGFR (Figures S15A-C). No CNV from the recipient nor the donor was associated with time-to-graft-failure (Figure S15D and E). However, a chromosome 2 duplication mismatch (HGSV_38325, tagged by the rs2392713 SNP, $r^2$=0.87 in KiT-GENIE) was significantly associated with time-to-kidney graft failure (Figure S15F and Table S6, p=1.1x$10^{-6}$, HR=4.5 [2.5-8.4], mismatch frequency=1.4%). The 20 pairs composed of a recipient not carrying the CNV while their donor exhibited the duplication had a 4-times increased risk of losing their kidney graft compared to the other pairs (Figure S16A). This duplication lies in an intergenic region between a block of genes 620kb upstream and the *EPHA4* gene 900kb downstream (Figure S16B). We tested the rs2392713 tagSNP for replication in the validation cohorts (Table S10, p=1.1x$10^{-6}$ in KiT-GENIE) but did not validate the signal (p=0.70).

Two deletion-tagging mismatches were previously reported associated with time-to-rejection under the genomic collision model: the *LIMS1* rs893403 mismatch (n=705 pairs)[11] and the *CFHR* rs7542235 mismatch (n=1,025 pairs)[29]. Both deletion mismatches were included in our CNV panel and were not associated with our outcomes of interest. Similarly to the original papers, we then tested them for association with time-to-rejection: the *LIMS1* deletion reached the multiple testing significance threshold in our cohort (p=0.001, HR=1.8 [1.3-2.6]) while the *CFHR* deletion did not reach nominal significance (p=0.69, Figure S17 and Table S11). Among the 39 tagSNPs from the original Steers *et al.* paper[11] present in our dataset, no other variant reached the Bonferroni significance threshold (p>0.0013, Table S11).

**Genome-wide polygenic scores |** Out of the 41,426 variants included in the most recent CKD GPS score[18], 39,490 (95%) were present in our KiT-GENIE dataset. Contrary to the original paper, we did not find any association between the score top 2% and the CKD status (p=0.28, same observation for top 1%, 5% and 10%, Table S12). Unlike the donor's CKD GPS, the recipients' CKD GPS was nominally associated with 1-year post-transplant eGFR (p=0.04, OR=2.7 [1.1-6.7], Table S13), but this effect was not sustained at 5 years (p=0.15). We did not observe any association between time-to-graft failure and the recipients' or the donors' CKD GPS (p=0.14 and p=0.57, respectively).

Regarding the genome-wide non-*HLA* mismatch score[10], 39,420 SNVs were included (*vs.* 59,268 in the original paper). As expected, the distribution of the overall mismatch count differed between related and unrelated pairs (Figure S19). We further dichotomized the scores in two groups (high and low according to the observed bimodal distribution) and found a Pearson correlation coefficient of R=-0.98 with the donor-recipient proportion of identity-by-descent value. The distribution of the mismatch count in unrelated pairs was similar to the original paper's (Figure S1), with a slightly lower median (1752 IQR 1712-1798 vs 1892 IQR 1850–1936). We implemented various iterations of the model, notably by restricting the analysis to pairs with deceased donors or excluding grafts with a rank 2 or higher, or by including additional SNVs (Table S15). We also computed a competing risk model, including death as a competing event. None of these analyses produced a nominally significant association (lowest p=0.08), and the observed effect's direction was almost systematically opposite to what was expected (HR<1, Table S15). In addition, we did not observe any association between the original non-*HLA* mismatch score and the post-transplant eGFR outcomes (p=0.94 at 1 year and p=0.42 at 5 years, Table S13), nor with time-to-rejection (p=0.19).

### Supplementary Tables

| | UKIRTC | Prague |
|---|---|---|
| Number of pairs | 1,477 | 365 |
| Median transplant year | 2001 | 2010 |
| Number of graft failure events | 364 (25%) | 54 (15%) |
| Percentage of biopsy-proven rejection events | 12% | 38% |
| Median HLA ABDR mismatches (/6) | 3 | 3 |
| Mean recipient age (years-old, sd) | 46.1 (14) | 42.2 (13) |
| Female recipient sex (n, %) | 554 (37%) | 131 (36%) |
| Mean donor age (years-old, sd) | 45.1 (14) | 49.3 (11) |
| Female donor sex (n, %) | 705 (48%) | 242 (66%) |
| Living donor (n, %) | 394 (27%) | 365 (100 %) |

**Supplementary Table S1 –** Demographic data for the two independent European validation cohorts

| | FAILURE | | |
|---|---|---|---|
| | **0**<br>N = 1,179[1] | **1**<br>N = 303[1] | **p-value**[2] |
| **Pre-transplant characteristics** | | | |
| Primary Disease | | | 3.1e-01 |
| Chronic glomerulonephritis | 324 (27%) | 91 (30%) | |
| Diabetes | 75 (6.4%) | 24 (7.9%) | |
| Tubulo-interstitial nephritis, deformities and others | 572 (49%) | 127 (42%) | |
| Unknown | 104 (8.8%) | 33 (11%) | |
| Vascular disease | 104 (8.8%) | 28 (9.2%) | |
| CMV infection (recipient) | 500 (43%) | 137 (45%) | 3.8e-01 |
| Unknown | 4 | 1 | |
| EBV infection (recipient) | 1,110 (95%) | 286 (96%) | 5.0e-01 |
| Unknown | 11 | 5 | |
| HCV infection (recipient) | 28 (2.4%) | 5 (1.7%) | 4.4e-01 |
| Unknown | 2 | 0 | |
| HIV infection (recipient) | 8 (0.7%) | 2 (0.7%) | 1.0e+00 |
| Unknown | 6 | 1 | |
| HIV infection (donor) | 1 (<0.1%) | 0 (0%) | 1.0e+00 |
| Unknown | 3 | 0 | |
| EBV infection (donor) | 1,128 (96%) | 284 (94%) | 1.8e-01 |
| Unknown | 2 | 1 | |
| CMV infection (donor) | 468 (40%) | 126 (42%) | 5.5e-01 |

[1] n (%); Median (Q1, Q3)

[2] Pearson's Chi-squared test; Fisher's exact test; Wilcoxon rank sum test

| | FAILURE | | |
|---|---|---|---|
| | **0**<br>N = 1,179[1] | **1**<br>N = 303[1] | **p-value**[2] |
| History of diabetes | 188 (16%) | 56 (18%) | 2.9e-01 |
| History of vascular disease | 299 (25%) | 76 (25%) | 9.2e-01 |
| History of neoplasia | 197 (17%) | 36 (12%) | 3.9e-02 |
| History of BK virus infection | 1 (<0.1%) | 1 (0.3%) | 3.7e-01 |
| History of hypertension | 1,074 (91%) | 279 (92%) | 5.9e-01 |
| History of squamous cell carcinoma | 16 (1.4%) | 5 (1.7%) | 7.8e-01 |
| History of basal cell carcinoma or Bowen's disease | 55 (4.7%) | 6 (2.0%) | 3.6e-02 |
| History of melanoma | 6 (0.5%) | 0 (0%) | 6.1e-01 |
| Preformed DSA | 515 (44%) | 139 (46%) | 4.9e-01 |
| Time preformed DSA (days) | -382 (-2,015, -151) | -720 (-3,260, -253) | 1.5e-02 |
| Unknown | 664 | 164 | |
| **Transplant characteristics** | | | |
| Graft rank | | | 6.3e-02 |
| 1 | 941 (80%) | 223 (74%) | |
| 2 | 200 (17%) | 67 (22%) | |
| 3 | 38 (3.2%) | 13 (4.3%) | |
| Graft year | 2012 (2008, 2016) | 2008 (2004, 2011) | **2.2e-28** |
| Recipient age | 54 (44, 64) | 55 (43, 65) | 5.4e-01 |
| Donor age | 55 (46, 65) | 57 (47, 70) | 1.9e-02 |

[1] n (%); Median (Q1, Q3)

[2] Pearson's Chi-squared test; Fisher's exact test; Wilcoxon rank sum test

| | FAILURE | | |
|---|---|---|---|
| | **0**<br>N = 1,179[1] | **1**<br>N = 303[1] | **p-value**[2] |
| Recipient sex | | | 3.8e-01 |
| F | 409 (35%) | 97 (32%) | |
| M | 770 (65%) | 206 (68%) | |
| Donor sex | | | 6.6e-01 |
| F | 495 (42%) | 123 (41%) | |
| M | 684 (58%) | 180 (59%) | |
| Induction treatment | | | 9.7e-01 |
| Depleting induction | 513 (44%) | 134 (44%) | |
| No induction | 27 (2.3%) | 7 (2.3%) | |
| Non-depleting induction | 639 (54%) | 162 (53%) | |
| CNI | 1,162 (99%) | 298 (98%) | 7.9e-01 |
| Treatment with a CNI-related agent | 16 (1.4%) | 2 (0.7%) | 5.5e-01 |
| Tacrolimus | 1,082 (92%) | 252 (83%) | **8.4e-06** |
| mTOR | 13 (1.1%) | 5 (1.7%) | 3.9e-01 |
| sirolimus | 2 (0.2%) | 3 (1.0%) | 6.1e-02 |
| everolimus | 11 (0.9%) | 2 (0.7%) | 1.0e+00 |
| Antiproliferation agent | 1,162 (99%) | 297 (98%) | 4.4e-01 |
| MMF | 745 (63%) | 265 (87%) | **6.1e-16** |
| MPA | 431 (37%) | 34 (11%) | **2.3e-17** |
| AZA | 10 (0.8%) | 2 (0.7%) | 1.0e+00 |
| Corticosteroids | 1,002 (85%) | 240 (79%) | 1.5e-02 |

[1] n (%); Median (Q1, Q3)

[2] Pearson’s Chi-squared test; Fisher’s exact test; Wilcoxon rank sum test

| | FAILURE | | |
|---|---|---|---|
| | **0**<br>N = 1,179[1] | **1**<br>N = 303[1] | **p-value**[2] |
| Donor blood related (up to 2nd degree) | 143 (12%) | 13 (4.3%) | **7.3e-05** |
| **Post-transplant characteristics** | | | |
| Resumption of function (days) | 3.0 (1.0, 8.0) | 4.0 (1.0, 12.0) | 9.7e-04 |
| Unknown | 6 | 7 | |
| Lost to follow-up | 103 (8.7%) | 13 (4.3%) | 1.0e-02 |
| Follow-up time (years) | 6.1 (3.9, 10.8) | 5.1 (2.3, 8.5) | **1.4e-07** |
| Post-Tx cardiac disease | 228 (19%) | 74 (24%) | 5.0e-02 |
| Post-Tx heart failure | 71 (6.0%) | 27 (8.9%) | 7.1e-02 |
| Post-Tx coronary insufficiency | 65 (5.5%) | 10 (3.3%) | 1.2e-01 |
| Post-Tx vascular disease | 191 (16%) | 43 (14%) | 3.9e-01 |
| Post-Tx cardiovascular disease | 347 (29%) | 99 (33%) | 2.7e-01 |
| Post-Tx hypertension | 176 (15%) | 62 (20%) | 1.9e-02 |
| Post-Tx arteriopathy | 67 (5.7%) | 11 (3.6%) | 1.5e-01 |
| Post-Tx stroke | 44 (3.7%) | 12 (4.0%) | 8.5e-01 |
| Post-Tx thromboembolic venous disease | 96 (8.1%) | 22 (7.3%) | 6.1e-01 |
| Post-Tx neoplasia | 310 (26%) | 55 (18%) | 3.3e-03 |
| Post-Tx basal cell carcinoma | 157 (13%) | 28 (9.2%) | 5.6e-02 |
| Post-Tx squamous cell carcinoma | 105 (8.9%) | 19 (6.3%) | 1.4e-01 |
| Post-Tx lymphoma | 14 (1.2%) | 6 (2.0%) | 2.7e-01 |
| De novo diabetes | 260 (22%) | 71 (23%) | 6.1e-01 |

[1] n (%); Median (Q1, Q3)

[2] Pearson's Chi-squared test; Fisher's exact test; Wilcoxon rank sum test

| | FAILURE | | |
|---|---|---|---|
| | **0**<br>N = 1,179[1] | **1**<br>N = 303[1] | **p-value**[2] |
| History of cardiac disease | 343 (29%) | 102 (34%) | 1.2e-01 |
| Bacterial infection | 619 (53%) | 190 (63%) | 1.5e-03 |
| Fungus infection | 58 (4.9%) | 14 (4.6%) | 8.3e-01 |
| Parasite infection | 16 (1.4%) | 1 (0.3%) | 2.2e-01 |
| Viral infection | 379 (32%) | 86 (28%) | 2.1e-01 |
| Urinary infection | 307 (26%) | 85 (28%) | 4.8e-01 |
| Pneumonia | 192 (16%) | 74 (24%) | 9.9e-04 |
| Pneumocytosis | 16 (1.4%) | 7 (2.3%) | 2.9e-01 |
| Sepsis | 94 (8.0%) | 46 (15%) | **1.3e-04** |
| Peritonitis | 12 (1.0%) | 1 (0.3%) | 4.9e-01 |
| Severe infection | 526 (45%) | 163 (54%) | 4.3e-03 |
| Primary disease recurrence | 32 (2.7%) | 26 (8.6%) | **2.6e-06** |
| De novo nephropathy | 16 (1.4%) | 6 (2.0%) | 4.3e-01 |
| BK virus nephropathy | 27 (2.3%) | 15 (5.0%) | 1.3e-02 |
| CNI toxicity | 30 (2.5%) | 19 (6.3%) | 1.2e-03 |
| At least one rejection event | 92 (7.8%) | 79 (26%) | **6.8e-19** |
| De novo DSA | 526 (45%) | 163 (54%) | 4.3e-03 |
| Time to de novo DSA (days) | 155 (79, 802) | 341 (44, 1,085) | 8.7e-01 |
| Unknown | 653 | 140 | |
| Proteinuria (at any post-Tx visit) | 969 (82%) | 268 (88%) | 8.9e-03 |

**Supplementary Table S2 –** Demographic table comparing the failure and control groups in KiT-GENIE for several non-genetic factors. *Tx, Transplantation.*

| | KIT-GENIE | UKIRTC | Prague |
|---|---|---|---|
| Alternative allele | A | A | A |
| Typed/Imputed ($R^2$) | Imputed (0.92) | Imputed (>0.8) | Imputed (0.88) |
| MISMATCH FREQ | 3.2% | 3.2% | 2.4% |
| RECIPIENT MAF | 2.3% | 2.0% | 1.1% |
| DONOR MAF | 1.8% | 1.8% | 1.2% |

**Supplementary Table S3 –** Frequencies for rs75606962, lead SNP of the chromosome 10 signal in the KiT-GENIE discovery cohort. *MAF, minor allele frequency.*

| | KIT-GENIE | UKIRTC | Prague |
|---|---|---|---|
| Alternative allele | A | A | A |
| Typed/Imputed ($R^2$) | Typed | Imputed (>0.8) | Typed |
| MISMATCH FREQ | 2.4% | 1.7% | 1.4% |
| RECIPIENT MAF | 1.5% | 1.1% | 1.0% |
| DONOR MAF | 1.3% | 0.9% | 1.3% |

**Supplementary Table S4 –** Frequencies for rs16955257, lead SNP of the chromosome 17 signal in the KiT-GENIE discovery cohort. *MAF, minor allele frequency.*

| | KIT-GENIE | UKIRTC | Prague |
|---|---|---|---|
| Alternative allele | G | G | G |
| Typed/Imputed ($R^2$) | Imputed (0.83) | Imputed (>0.8) | Imputed (0.92) |
| MISMATCH FREQ | 1.7% | 3.4% | 3.6% |
| RECIPIENT MAF | 1.3% | 1.3% | 2.2% |
| DONOR MAF | 0.8% | 1.7% | 2.1% |

**Supplementary Table S5 –** Frequencies for rs117176234, lead SNP of the chromosome 21 signal in the KiT-GENIE discovery cohort. *MAF, minor allele frequency.*

| Variant ID | CHR | $P_{discovery}$ | $HR_{discovery}$ | $P_{meta-analysis-VAL}$ | $HR_{meta-analysis-VAL}$ | $p_{heterogeneity VAL}$ |
|---|---|---|---|---|---|---|
| HGSV_38325 | 2 | $1.1.x10^{-6}$ | 4.5 [2.5-8.4] | NA | NA | NA |
| rs2392713 (tagSNP for HGSV_38325) | 2 | $1.1x10^{-6}$ | 4.1 [2.2 – 7.5] | 0.70 | 0.9 [0.5 – 1.5] | 0.90 |
| rs75606962 | 10 | $2.0x10^{-8}$ | 3.5 [2.3-5.4] | 0.48 | 1.2 [0.7-2.0] | 0.99 |
| **rs16955257** | **17** | **$6.3x10^{-9}$** | **4.1 [2.5-6.5]** | **0.02** | **1.9 [1.4-3.2]** | 0.29 |
| rs117176234 | 21 | $3.6x10^{-8}$ | 5.1 [2.9-9.0] | 0.70 | 0.9 [0.5-1.5] | 0.37 |

**Supplementary Table S6 –** GWSS summary statistics for the SNP and CNV mismatches significantly associated with time-to-graft failure in the original KiT-GENIE cohort, and in the meta-analysis across the two validation cohorts. The chr2 CNV data were not available in the validation cohorts, so replication was assessed with the rs2392713 tagSNP (LD of $r^2$=0.87 in KiT-GENIE). The replicated chr17 SNP mismatch is highlighted in bold. Chr, chromosome; HR, hazards-ratio; VAL, validation cohorts.

| **Model** | **P** | **HR** |
|---|---|---|
| Original paper summary statistics*(n=1,482, 303 events) | $6.3x10^{-9}$ | 4.1 [2.6-6.5] |
| Original model with mismatches computed as the absolute D-R difference (n=1,482, 303 events) | $1.3x10^{-4}$ | 2.2 [1.5-3.4] |
| Original model including rejection events in covariates (n=1,482, 303 events) | $9.6x10^{-10}$ | 4.4 [2.7-7.0] |
| Original model with primary disease recurrence as covariate (n=1,482, 303 events) | $3.7x10^{-8}$ | 3.8 [2.4-6.1] |
| Original model with tacrolimus at induction (yes/no) as covariate (n=1,482, 303 events) | $1.4x10^{-8}$ | 4.0 [2.5-6.4] |
| Original model with MMF at induction (yes/no) as covariate (n=1,482, 303 events) | $1.2x10^{-8}$ | 4.0 [2.5-6.4] |
| Original model by adding a level to donor status to consider relatedness (n=1,482, 303 events) | $6.3x10^{-9}$ | 4.1 [2.6-6.5] |
| Original model with the Euclidian distance in PC1xPC2 space as covariate (n=1,482, 303 events) | $1.0x10^{-8}$ | 4.0 [2.5-6.4] |
| Original model considering relatedness and the six above additional covariates, n=1,482, 303 events) | $3.1x10^{-8}$ | 3.9 [2.4-6.2] |
| Association with time-to-rejection (n=1,482, 171 events) | 0.62 | 1.3 [0.5-3.5] |
| Association with time-to-ABMR (n=1,482, 85 events) | 0.11 | 2.6 [0.8-8.3] |
| Association with time-to-TCMR (n=1,482, 113 events) | 0.62 | 1.3 [0.4-4.3] |
| Sensitivity analysis in Tx achieving primary graft function – association with time-to-GF (n=950, 159 events) | $3.9x10^{-5}$ | 4.3 [2.1-8.5] |
| Sensitivity analysis in Tx achieving primary graft function – association with time-to-ABMR (n=950, 58 events) | 0.02 | 4.1 [1.3-13.8] |
| Sensitivity analysis in Tx achieving primary graft function – association with time-to-TCMR (n=950, 63 events) | 0.02 | 4.2 [1.3-13.8] |

| | | |
|---|---|---|
| Competing risk model (death with functioning graft as a competing event, n=1,482) | $1.1x10^{-8}$ | 4.1 [2.5-6.6] |

**Supplementary Table S7** – Assessment of the replicated *TOM1L1* signal (rs16955257) robustness across various models, adjustments and subgroups from the KiT-GENIE discovery cohort. The summary statistics were obtained in the discovery cohort exclusively. *The original model is a Cox proportional hazards model fitted for time-to-failure and adjusted for graft year, graft rank, recipient and donor age and sex, HLA D-R epitopic mismatches, donor and recipient first four PCs. *Tx, Transplantation. GF, Graft Failure. ABMR, Antibody mediated rejection. TCMR, T-cell mediated rejection*.

| Cause for allograft failure | rs16955257 | | p- value[2] |
|---|---|---|---|
| | MATCH N =284[1] | MISMATCH N = 19[1] | |
| Graft failure cause | | | **4.6e-02** |
| BK virus infection | 8 (2.8%) | 0 (0%) | |
| Chronic graft dysfunction | 148 (52%) | 6 (32%) | |
| Chronic humoral rejection | 19 (6.7%) | 0 (0%) | |
| Disease recurrence | 18 (6.3%) | 2 (11%) | |
| Graft infection | 6 (2.1%) | 1 (5.3%) | |
| Hyperacute vascular rejection | 3 (1.1%) | 0 (0%) | |
| Immunosuppression treatment interruption due to medical cause | 2 (0.7%) | 1 (5.3%) | |
| Rejection after immunosuppression treatment interruption | 7 (2.5%) | 0 (0%) | |
| Rejection under immunosuppression treatment | 14 (4.9%) | 2 (11%) | |
| Urologic cause | 2 (0.7%) | 0 (0%) | |
| Vascular cause | 11 (3.9%) | 4 (21%) | |
| Other cause | 9 (3.2%) | 1 (5.3%) | |
| Unknown | 37 (13%) | 2 (11%) | |

[1] n (%)

[2] Fisher's exact test

**Supplementary Table S8** – Comparison of the reported graft failure cause between the *TOM1L1* chr17 matched and mismatched groups in the KiT-GENIE discovery cohort. A nominally significant difference was observed between the matched and the mismatched group, with a higher rate of failure events caused by disease recurrence and vascular disease in the mismatched group. We want to call for cautious in interpreting these results considering the low numbers of events for each cause ($1 \leq n \leq 6$) in the chr17 mismatched group.

| | rs16955257 | | |
|---|---|---|---|
| | **MATCH** N = 1,446[1] | **MISMATCH** N = 36[1] | **p-value**[2] |
| **Pre-transplant characteristics** | | | |
| Primary Disease | | | 7.6e-01 |
| Chronic glomerulonephritis | 402 (28%) | 13 (36%) | |
| Diabetes | 96 (6.6%) | 3 (8.3%) | |
| Tubulo-interstitial nephritis, deformities and others | 685 (47%) | 14 (39%) | |
| Unknown | 134 (9.3%) | 3 (8.3%) | |
| Vascular disease | 129 (8.9%) | 3 (8.3%) | |
| CMV infection (recipient) | 622 (43%) | 15 (42%) | 8.6e-01 |
| Unknown | 5 | 0 | |
| EBV infection (recipient) | 1,361 (95%) | 35 (97%) | 1.0e+00 |
| Unknown | 16 | 0 | |
| HCV infection (recipient) | 33 (2.3%) | 0 (0%) | 1.0e+00 |
| Unknown | 2 | 0 | |
| HIV infection (recipient) | 10 (0.7%) | 0 (0%) | 1.0e+00 |
| Unknown | 7 | 0 | |
| HIV infection (donor) | 1 (<0.1%) | 0 (0%) | 1.0e+00 |
| Unknown | 3 | 0 | |
| EBV infection (donor) | 1,377 (95%) | 35 (97%) | 1.0e+00 |
| Unknown | 3 | 0 | |
| CMV infection (donor) | 580 (40%) | 14 (39%) | 8.8e-01 |

[1] n (%); Median (Q1, Q3)

[2] Fisher's exact test; Pearson's Chi-squared test; Wilcoxon rank sum test

| | rs16955257 | | |
|---|---|---|---|
| | **MATCH**<br>N = 1,446[1] | **MISMATCH**<br>N = 36[1] | **p-value**[2] |
| History of diabetes | 240 (17%) | 4 (11%) | 3.8e-01 |
| History of vascular disease | 366 (25%) | 9 (25%) | 9.7e-01 |
| History of neoplasia | 230 (16%) | 3 (8.3%) | 2.2e-01 |
| History of BK virus infection | 2 (0.1%) | 0 (0%) | 1.0e+00 |
| History of hypertension | 1,322 (91%) | 31 (86%) | 2.3e-01 |
| History of squamous cell carcinoma | 21 (1.5%) | 0 (0%) | 1.0e+00 |
| History of basal cell carcinoma or Bowen's disease | 59 (4.1%) | 2 (5.6%) | 6.6e-01 |
| History of melanoma | 6 (0.4%) | 0 (0%) | 1.0e+00 |
| Preformed DSA | 636 (44%) | 18 (50%) | 4.7e-01 |
| Time preformed DSA (days) | -451 (-2,249, -156) | -700 (-920, -163) | 9.7e-01 |
| Unknown | 810 | 18 | |
| **Transplant characteristics** | | | |
| Graft rank | | | 4.8e-01 |
| 1 | 1,135 (78%) | 29 (81%) | |
| 2 | 262 (18%) | 5 (14%) | |
| 3 | 49 (3.4%) | 2 (5.6%) | |
| Graft year | 2011 (2007, 2015) | 2008 (2005, 2013) | 1.3e-02 |
| Recipient age | 54 (44, 64) | 58 (42, 67) | 3.4e-01 |
| Donor age | 55 (46, 65) | 58 (50, 72) | 1.5e-01 |

[1] n (%); Median (Q1, Q3)

[2] Fisher's exact test; Pearson's Chi-squared test; Wilcoxon rank sum test

| | rs16955257 | | |
|---|---|---|---|
| | **MATCH** N = 1,446[1] | **MISMATCH** N = 36[1] | **p-value**[2] |
| Recipient sex | | | 8.0e-01 |
| F | 493 (34%) | 13 (36%) | |
| M | 953 (66%) | 23 (64%) | |
| Donor sex | | | 1.7e-01 |
| F | 607 (42%) | 11 (31%) | |
| M | 839 (58%) | 25 (69%) | |
| Induction treatment | | | 1.7e-02 |
| Depleting induction | 633 (44%) | 14 (39%) | |
| No induction | 30 (2.1%) | 4 (11%) | |
| Non-depleting induction | 783 (54%) | 18 (50%) | |
| CNI | 1,426 (99%) | 34 (94%) | 9.8e-02 |
| Treatment with a CNI-related agent | 18 (1.2%) | 0 (0%) | 1.0e+00 |
| Tacrolimus | 1,308 (90%) | 26 (72%) | 1.9e-03 |
| mTOR | 17 (1.2%) | 1 (2.8%) | 3.6e-01 |
| sirolimus | 5 (0.3%) | 0 (0%) | 1.0e+00 |
| everolimus | 12 (0.8%) | 1 (2.8%) | 2.7e-01 |
| Antiproliferation agent | 1,423 (98%) | 36 (100%) | 1.0e+00 |
| MMF | 979 (68%) | 31 (86%) | 1.9e-02 |
| MPA | 460 (32%) | 5 (14%) | 2.2e-02 |
| AZA | 12 (0.8%) | 0 (0%) | 1.0e+00 |
| Corticosteroids | 1,213 (84%) | 29 (81%) | 5.9e-01 |

[1] n (%); Median (Q1, Q3)

[2] Fisher's exact test; Pearson's Chi-squared test; Wilcoxon rank sum test

| | rs16955257 | | |
|---|---|---|---|
| | **MATCH** N = 1,446[1] | **MISMATCH** N = 36[1] | **p-value**[2] |
| Blood related donor (up to 2nd degree) | 154 (11%) | 2 (5.6%) | 5.8e-01 |
| **Post-transplant characteristics** | | | |
| Resumption of function (days) | 3.0 (1.0, 8.0) | 4.0 (2.0, 8.0) | 4.7e-01 |
| Unknown | 11 | 2 | |
| Lost to follow-up | 113 (7.8%) | 3 (8.3%) | 7.6e-01 |
| Follow-up time (years) | 6.0 (3.3, 10.1) | 4.0 (1.2, 8.1) | 2.8e-03 |
| Post-Tx cardiac disease | 297 (21%) | 5 (14%) | 3.3e-01 |
| Post-Tx heart failure | 98 (6.8%) | 0 (0%) | 1.7e-01 |
| Post-Tx coronary insufficiency | 75 (5.2%) | 0 (0%) | 2.6e-01 |
| Post-Tx vascular disease | 225 (16%) | 9 (25%) | 1.2e-01 |
| Post-Tx cardiovascular disease | 434 (30%) | 12 (33%) | 6.7e-01 |
| Post-Tx hypertension | 228 (16%) | 10 (28%) | 5.3e-02 |
| Post-Tx arteriopathy | 77 (5.3%) | 1 (2.8%) | 1.0e+00 |
| Post-Tx stroke | 54 (3.7%) | 2 (5.6%) | 6.4e-01 |
| Post-Tx thromboembolic venous disease | 112 (7.7%) | 6 (17%) | 6.1e-02 |
| Post-Tx neoplasia | 360 (25%) | 5 (14%) | 1.3e-01 |
| Post-Tx basal cell carcinoma | 183 (13%) | 2 (5.6%) | 3.0e-01 |
| Post-Tx squamous cell carcinoma | 123 (8.5%) | 1 (2.8%) | 3.6e-01 |
| Post-Tx lymphoma | 19 (1.3%) | 1 (2.8%) | 3.9e-01 |
| De novo diabetes | 324 (22%) | 7 (19%) | 6.7e-01 |

[1] n (%); Median (Q1, Q3)

[2] Fisher's exact test; Pearson's Chi-squared test; Wilcoxon rank sum test

| | rs16955257 | | |
|---|---|---|---|
| | **MATCH**<br>N = 1,446[1] | **MISMATCH**<br>N = 36[1] | **p-value**[2] |
| History of cardiac disease | 430 (30%) | 15 (42%) | 1.2e-01 |
| Bacterial infection | 790 (55%) | 19 (53%) | 8.3e-01 |
| Fungus infection | 70 (4.8%) | 2 (5.6%) | 6.9e-01 |
| Parasite infection | 17 (1.2%) | 0 (0%) | 1.0e+00 |
| Viral infection | 460 (32%) | 5 (14%) | 2.2e-02 |
| Urinary infection | 384 (27%) | 8 (22%) | 5.6e-01 |
| Pneumonia | 260 (18%) | 6 (17%) | 8.4e-01 |
| Pneumocytosis | 23 (1.6%) | 0 (0%) | 1.0e+00 |
| Sepsis | 134 (9.3%) | 6 (17%) | 1.4e-01 |
| Peritonitis | 12 (0.8%) | 1 (2.8%) | 2.7e-01 |
| Severe infection | 673 (47%) | 16 (44%) | 8.0e-01 |
| Primary disease recurrence | 55 (3.8%) | 3 (8.3%) | 1.6e-01 |
| De novo nephropathy | 21 (1.5%) | 1 (2.8%) | 4.2e-01 |
| BK virus nephropathy | 41 (2.8%) | 1 (2.8%) | 1.0e+00 |
| CNI toxicity | 48 (3.3%) | 1 (2.8%) | 1.0e+00 |
| At least one rejection event | 167 (12%) | 4 (11%) | 1.0e+00 |
| De novo DSA | 676 (47%) | 13 (36%) | 2.1e-01 |
| Time to de novo DSA (days) | 180 (77, 1,029) | 99 (84, 1,470) | 9.6e-01 |
| Unknown | 770 | 23 | |
| Proteinuria (at anly post-Tx visit) | 1,210 (84%) | 27 (75%) | 1.7e-01 |

[1] n (%); Median (Q1, Q3)

[2] Fisher's exact test; Pearson's Chi-squared test; Wilcoxon rank sum test

**Supplementary Table S9** - Demographic table comparing the chromosome 17 matched and mismatched groups for the replicated lead *TOM1L1* SNP signal (rs16955257) in KiT-GENIE for several non-genetic factors. When accounting for multiple testing ($P<7.1x10^{-4}$), no significant difference was observed. *Tx, transplantation.*

| | KIT-GENIE | UKIRTC | Prague |
|---|---|---|---|
| Alternative allele | G | G | G |
| Typed/Imputed ($R^2$) | Typed | Typed | Typed |
| MISMATCH FREQ | 1.4% | 1.0% | 1.1% |
| RECIPIENT MAF | 10.6% | 9.8% | 7.9% |
| DONOR MAF | 11.2% | 10.8% | 7.5% |

**Supplementary Table S10 –** Frequencies for rs2392713, tagSNP for the chromosome 2 CNV signal in the KiT-GENIE discovery cohort.

| Tagging SNP | CHR | A1 | A2 | A1 model | | | A2 model | | |
|---|---|---|---|---|---|---|---|---|---|
| | | | | Control (N=1371)* | Rejection (N=111)* | pvalue | Control (N=1371)* | Rejection (N=111)* | pvalue |
| rs10927864 | chr1 | C | G | 0.22 | 0.2 | 0.7705 | 0.11 | 0.06 | 0.075 |
| rs11249248 | chr1 | T | C | 0.15 | 0.15 | 0.8184 | 0.21 | 0.2 | 0.9189 |
| rs11209948 | chr1 | G | T | 0.11 | 0.1 | 0.6689 | 0.22 | 0.25 | 0.3989 |
| rs6693105 | chr1 | T | C | 0.09 | 0.05 | 0.0548 | 0.24 | 0.21 | 0.3378 |
| rs11587012 | chr1 | A | C | 0.22 | 0.28 | 0.1032 | 0.1 | 0.09 | 0.8112 |
| rs7542235 | chr1 | A | G | 0.23 | 0.23 | 0.9247 | 0.04 | 0.05 | 0.6851 |
| rs158736 | chr1 | G | C | 0.24 | 0.29 | 0.163 | 0.07 | 0.09 | 0.4171 |
| **rs893403** | **chr2** | **G** | **A** | **0.14** | **0.24** | **0.001** | 0.2 | 0.16 | 0.1787 |
| rs7419565 | chr2 | T | C | 0.21 | 0.25 | 0.5867 | 0.12 | 0.08 | 0.1196 |
| rs7703761 | chr5 | C | T | 0.15 | 0.15 | 0.909 | 0.21 | 0.2 | 0.8313 |
| rs10053292 | chr5 | T | C | 0.19 | 0.17 | 0.6591 | 0.01 | 0.02 | 0.4588 |
| rs2387715 | chr5 | A | T | 0.23 | 0.24 | 0.9295 | 0.08 | 0.11 | 0.1788 |
| rs17654108 | chr6 | A | T | 0.19 | 0.14 | 0.1518 | 0.02 | 0.01 | 0.2336 |
| rs2160195 | chr7 | A | T | 0.22 | 0.22 | 0.8175 | 0.05 | 0.05 | 0.9842 |
| rs4729606 | chr7 | T | C | 0.23 | 0.25 | 0.6914 | 0.05 | 0.04 | 0.2738 |
| rs6943474 | chr7 | A | G | 0.18 | 0.2 | 0.5597 | 0.2 | 0.2 | 0.8742 |
| rs4621754 | chr7 | A | G | 0.15 | 0.13 | 0.7597 | 0.02 | 0.01 | 0.2821 |
| rs4543566 | chr8 | C | G | 0.17 | 0.15 | 0.6432 | 0.02 | 0.01 | 0.594 |
| rs11985201 | chr8 | G | A | 0.2 | 0.21 | 0.9618 | 0.15 | 0.18 | 0.4818 |
| rs1523688 | chr9 | T | G | 0.23 | 0.18 | 0.0758 | 0.04 | 0.05 | 0.6421 |
| rs2174926 | chr9 | A | G | 0.15 | 0.19 | 0.1941 | 0.2 | 0.15 | 0.0878 |
| rs2342606 | chr10 | T | C | 0.16 | 0.17 | 0.8142 | 0.19 | 0.19 | 0.7711 |
| rs10885336 | chr10 | G | A | 0.23 | 0.2 | 0.6048 | 0.11 | 0.11 | 0.8945 |
| rs3793917 | chr10 | C | G | 0.23 | 0.26 | 0.414 | 0.06 | 0.04 | 0.2912 |
| rs4882017 | chr11 | A | G | 0.12 | 0.17 | 0.1351 | 0.22 | 0.22 | 0.9928 |
| rs11228868 | chr11 | C | T | 0.15 | 0.12 | 0.3198 | 0.01 | 0.01 | 0.7462 |

| | | | | | | | | | |
|---|---|---|---|---|---|---|---|---|---|
| rs1944862 | chr11 | G | A | 0.24 | 0.24 | 0.8077 | 0.07 | 0.05 | 0.9145 |
| rs1478309 | chr12 | T | G | 0.05 | 0.05 | 0.9482 | 0.2 | 0.3 | 0.0063 |
| rs9318648 | chr13 | A | G | 0.06 | 0.06 | 0.9095 | 0.24 | 0.22 | 0.5288 |
| rs8007442 | chr14 | T | C | 0.12 | 0.08 | 0.0784 | 0.22 | 0.21 | 0.9293 |
| rs11156875 | chr14 | A | G | 0.18 | 0.19 | 0.9565 | 0.02 | 0.04 | 0.4312 |
| rs8022070 | chr14 | C | T | 0.16 | 0.16 | 0.751 | 0.02 | 0.03 | 0.4547 |
| rs8025963 | chr15 | A | T | 0.02 | 0.05 | 0.035 | 0 | 0 | NA |
| rs10521145 | chr16 | G | A | 0.18 | 0.2 | 0.5277 | 0.02 | 0.01 | 0.6031 |
| rs16966699 | chr17 | C | G | 0.23 | 0.21 | 0.4057 | 0.05 | 0.03 | 0.2179 |
| rs4806152 | chr19 | A | C | 0.23 | 0.23 | 0.7741 | 0.04 | 0.05 | 0.3575 |
| rs324121 | chr19 | G | A | 0.17 | 0.18 | 0.7995 | 0.02 | 0.01 | 0.7142 |
| rs103294 | chr19 | T | C | 0.02 | 0.02 | 0.9135 | 0.22 | 0.2 | 0.6353 |
| rs3810336 | chr19 | G | A | 0.22 | 0.25 | 0.5523 | 0.09 | 0.14 | 0.0707 |

*Mismatch frequency

**Supplementary Table S11 –** Genomic collision model replication for CNV tagging SNPs in the KiT-GENIE cohort. Results of the association tests between the high-priority deletion tagging SNPs selected by *Steers et al*. and time-to-rejection (39 of the 44 SNPs were present in KiT-GENIE). Mismatches were computed in two different ways: A1 homozygous recipient with A1 non-homozygous donors (columns 5-7), and A2 homozygous recipient with A2 non-homozygous donors (columns 8-10). The *LIMS1* deletion tagging SNP (rs893403, in bold), reached the adjusted Bonferroni threshold (p=0.001, HR=1.8 [1.3-2.6]), while the *CFHR* deletion tagging SNP (rs7542235, p=0.7) did not reach nominal significance.

| | N in top x% | P-value | Effect size |
|---|---|---|---|
| Overall | NA | 0.68 | -0.02 [-0.09;0.06] |
| Top 10% | 303 | 0.25 | -0.14 [-0.39; 0.10] |
| Top 5% | 152 | 0.26 | -0.19 [-0.53;0.14] |
| Top 2% | 61 | 0.28 | 0.29 [-0.28 ;0.86] |
| Top 1% | 31 | 0.92 | 0.04 [-0.69;0.76] |

**Supplementary Table S12 –** Association between the CKD GPS (as proposed by Khan *et al.*), as well as its top quantiles, and the CKD outcome (assessed by comparing recipients vs donors) in the KiT-GENIE cohort.

| Score | **Time-to-graft failure** | **1-year eGFR** | **5-year eGFR** |
|---|---|---|---|
| Recipient's CKD GPS [1] | p=0.14<br>HR=0.8 [0.5-1.1] | p=0.04<br>OR=2.7 [1.1-6.7] | p=0.15<br>OR=2.5 [0.7-8.7] |
| Donor's CKD GPS [1] | p=0.57<br>HR=0.9 [0.6 – 1<br>.3] | p=0.05<br>OR=2.5 [1.0-6.5] | p=0.07<br>OR=3.3 [0.9-11.9] |
| Non-HLA [2] | p=0.14<br>HR=0.9 [0.8-1.0] | p=0.94<br>OR=1.0 [0.6-1.7] | p=0.42<br>OR=1.4 [0.6 – 0.3] |
| HLA ABDR (/6) | p=0.01<br>HR=1.10 [1.03-1.19] | p=0.18<br>OR=1.5 [0.8 – 2.7] | p=0.74<br>1.4 [0.6 – 3.0] |
| HLA class II epitopic mismatches | p=0.01<br>HR=1.2 [1.0-1.3] | p=0.5<br>OR=1.0 [0.9 -1.0] | p=0.5<br>OR=1.0 [0.9 – 1.1] |

**Supplementary Table S13 –** Polygenic risk scores summary statistics in the KiT-GENIE cohort. Association p-values between several scores vs. time-to-graft failure, 1-year post-transplant eGFR and 5-year post-transplant eGFR. [1] As proposed by Khan et al.; [2] As proposed by Reindl-Schwaighofer et al.

| | **CKD GPS** | **Non-HLA score** | ***LIMS1* tagSNP** | ***CFHR* tagSNP** |
|---|---|---|---|---|
| Outcome | Kidney failure | Kidney graft failure | Rejection | Rejection |
| Number of samples | 3,011 | 1,482 | 1,482 | 1,482 |
| Number of events | 1,687 | 303 | 171 | 171 |
| Effect size | 2.3 | 1.7 | 1.55 | 3.1 |
| Power | 100% | 100% | 67% | 86% |

**Supplementary Table S14** - Power analyses' results for replication of literature results tested in KiT-GENIE (alpha=5%).

| | Number of samples | Number of SNVs | P-value | HR |
|---|---|---|---|---|
| Main model* | 1,482 | 39,420 | 0.14 | 0.93 [0.84-1.03] |
| Association with score quartiles | 1,482 | 39,420 | 0.34 | 0.95 [0.85 – 1.06] |
| All SNVs | 1,482 | 22,463,451 | 0.08 | 1.19 [1.02 – 1.39] |
| All coding SNVs | 1,482 | 59,268 | 0.52 | 0.98 [0.93 – 1.04] |
| Main model restricted to deceased donors only | 1,207 | 39,420 | 0.10 | 0.86 [0.72-1.03] |
| Main model restricted to graft rank 1 only | 1,164 | 39,420 | 0.08 | 0.91 [0.82 – 1.01] |
| Competing events model (with death with a functioning graft as competing event) | 1,482 | 39,420 | 0.13 | 0.93 [0.84 – 1.02] |
| Association with time-to-rejection (171 events) | 1,482 | 39,420 | 0.19 | 1.00 [1.00-1.00] |

**Supplementary Table S15** – Summary statistics of the non-HLA mismatch score association with time-to-graft failure under various configurations. *The main model captures the association between the non-HLA genetic mismatch score (focusing on non-synonymous SNVs within genes encoding secreted and transmembrane proteins) and time-to-graft failure with a Cox proportional hazard model adjusted for donor age and sex, donor type (living or deceased) and HLA epitopic mismatches.

### Supplementary Figures

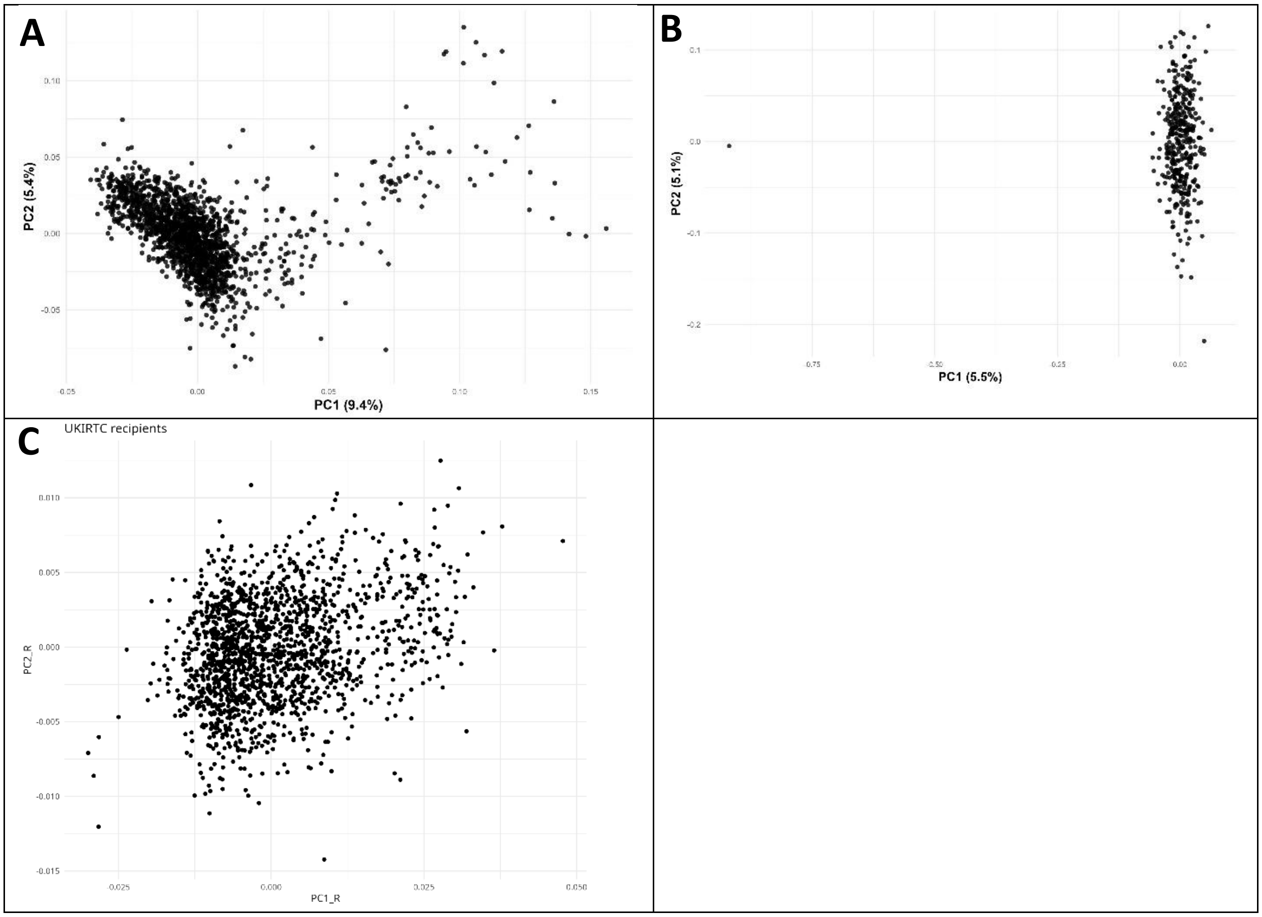

**Supplementary Figure S1** - PCA plots for recipients in the KiT-GENIE discovery cohort (A) and the replication cohorts (B=Prague, C=UKIRTC).

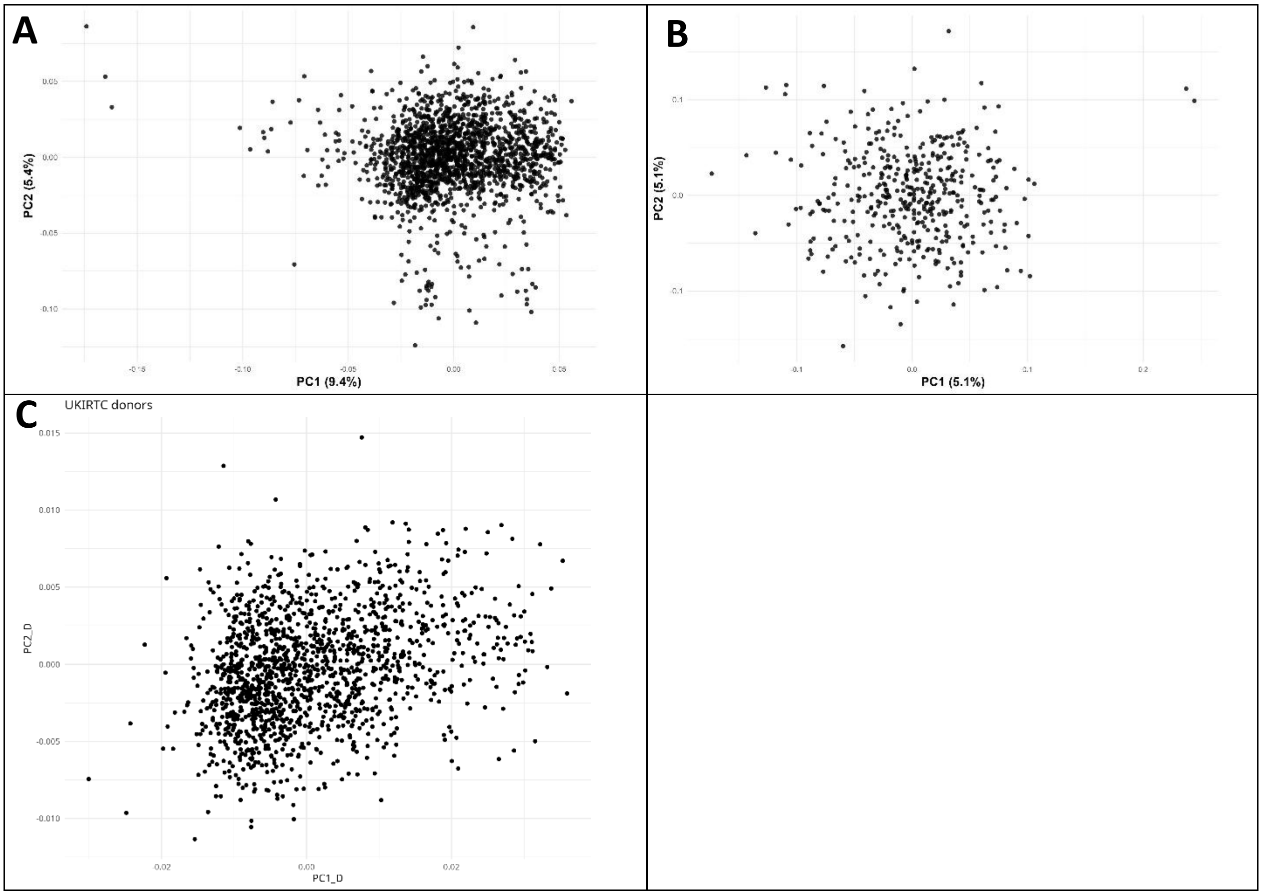

**Supplementary Figure S2 -** PCA plots for donors in the KiT-GENIE discovery cohort (A) and the replication cohorts (B=Prague, C=UKIRTC).

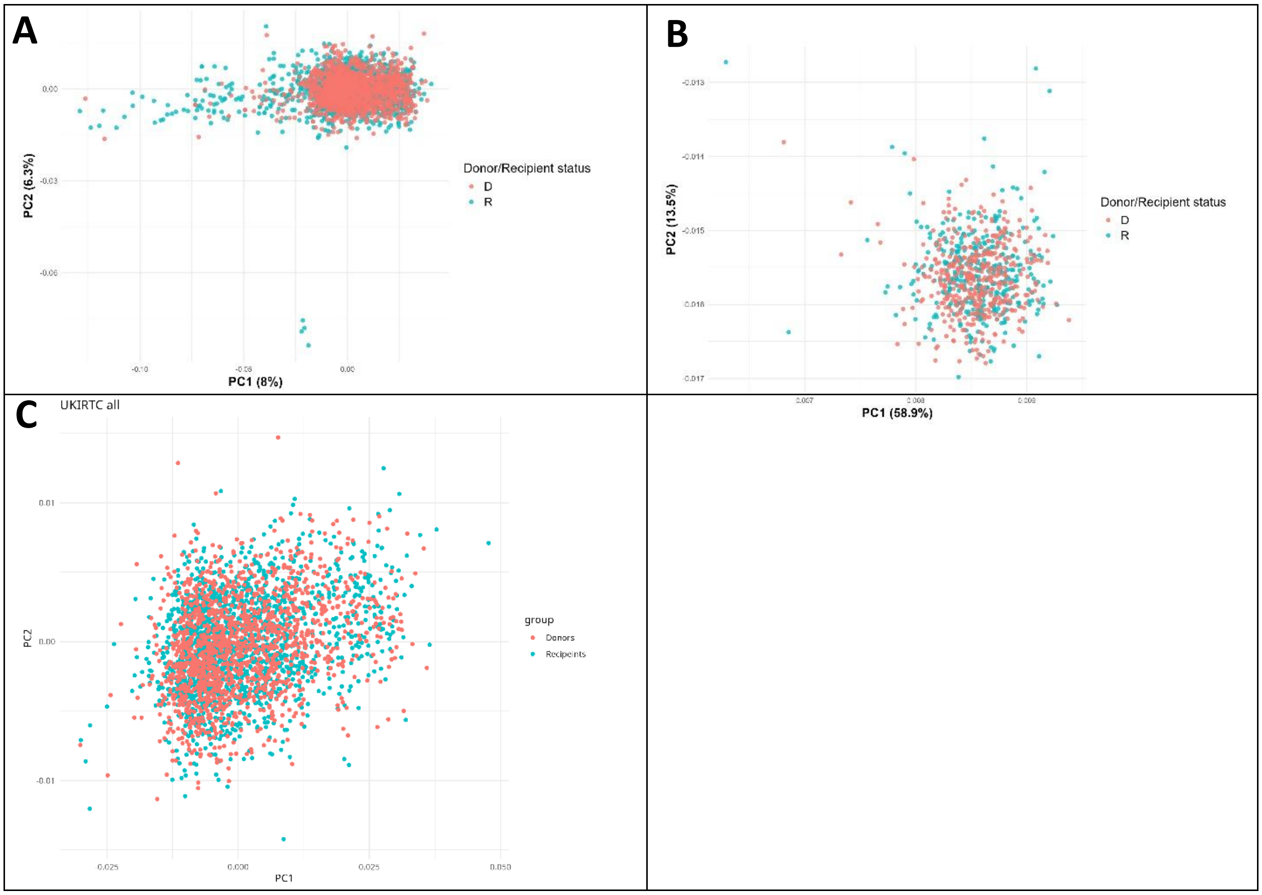

**Supplementary Figure S3 -** PCA plots for donors and recipients simultaneously in the KiT-GENIE discovery cohort (A) and the replication cohorts (B=Prague, C=UKIRTC).

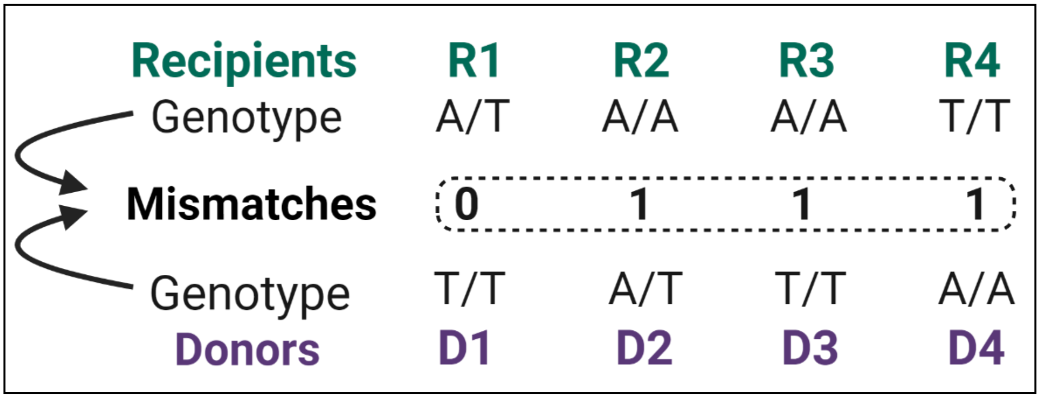

**Supplementary Figure S4 –** Illustration of donor-recipient mismatches computation principle with two alleles A and T. A mismatch was defined as an allele present in the donor but not in the recipient (equivalent to a 0/1 dominant model).

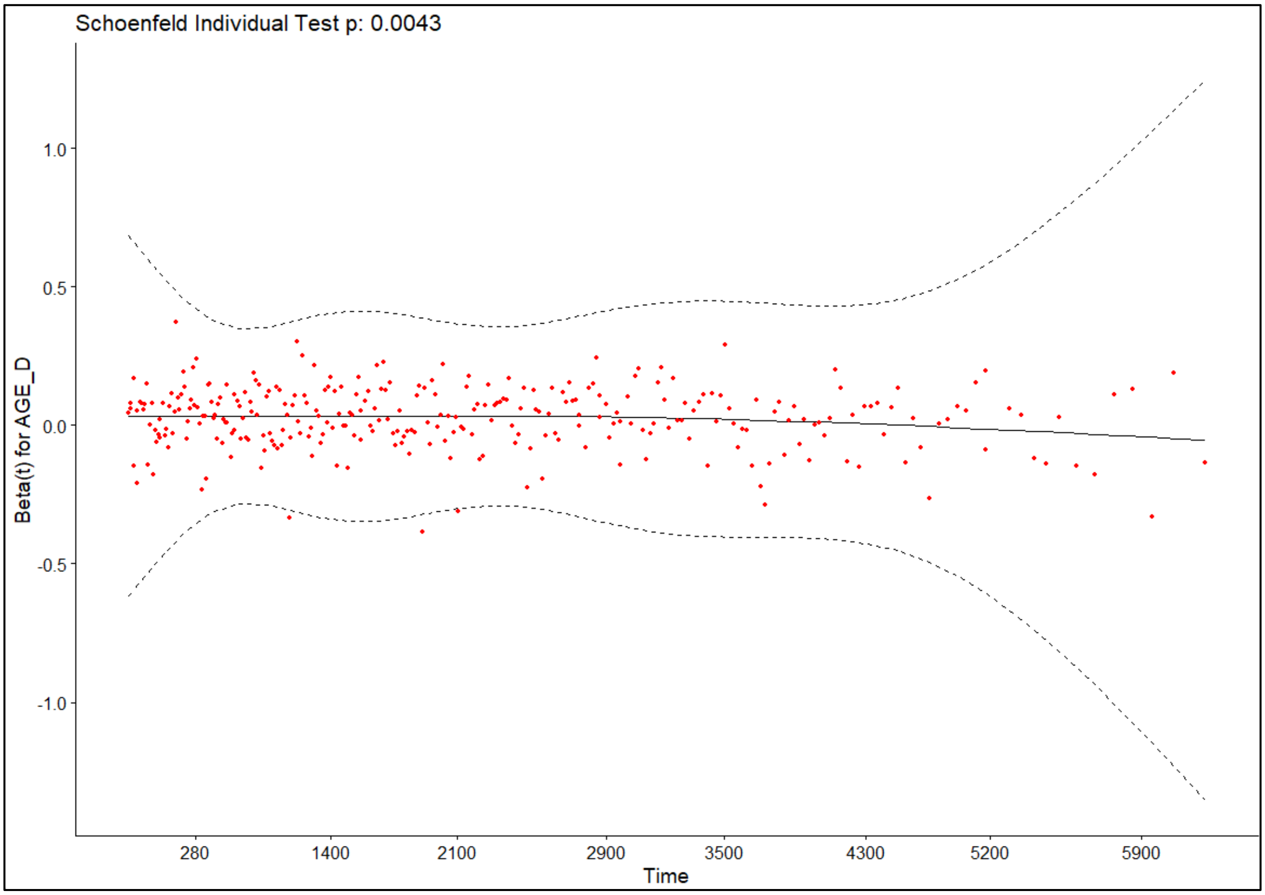

**Supplementary Figure S5** - Proportional hazards Cox model diagnostic - Schoenfeld residuals for donor age slowly decreased over time in KiT-GENIE donor-recipient pairs.

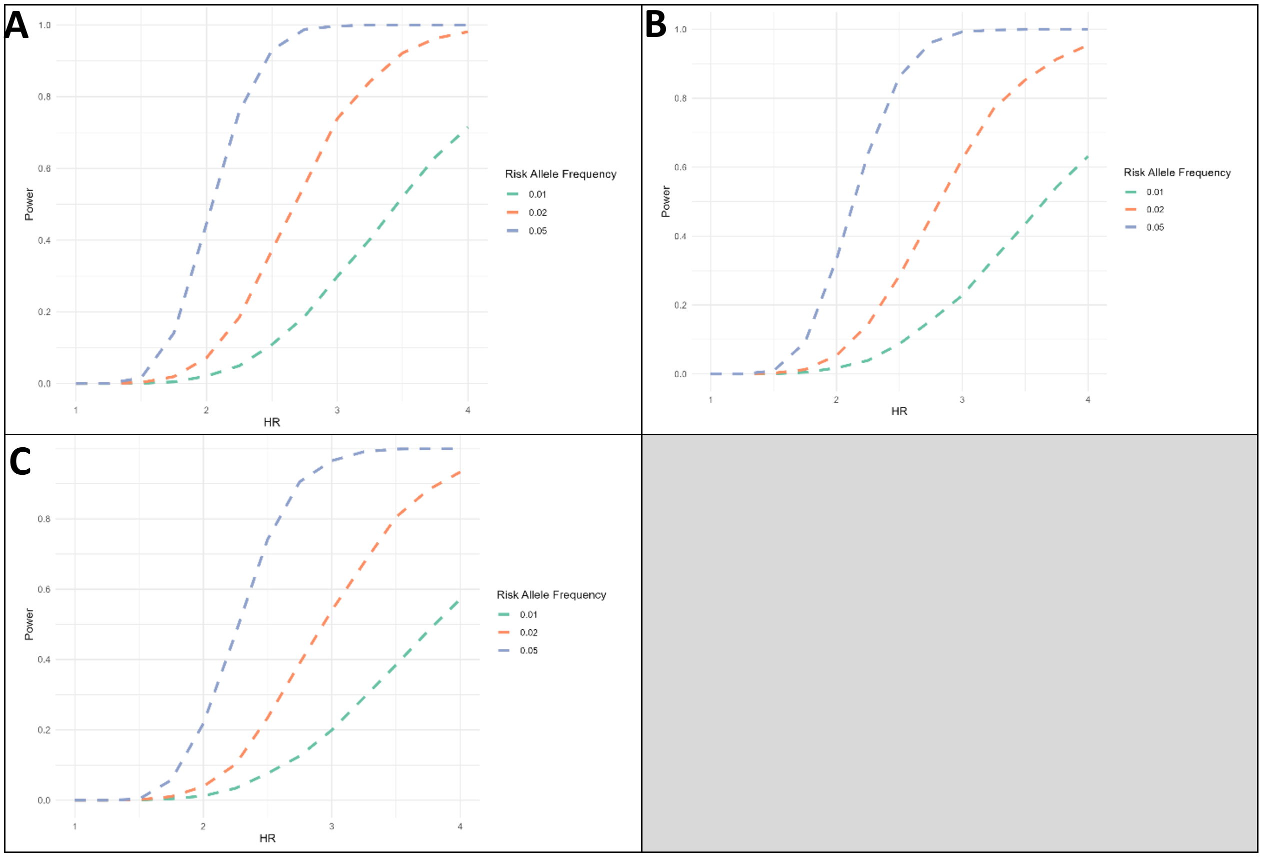

**Supplementary Figure S6 –** Statistical power analysis curves for the time-to-graft failure GWSS in recipients (A, additive model), donors (B, additive model) and pairs (C, dominant model) for the KiT-GENIE discovery cohort.

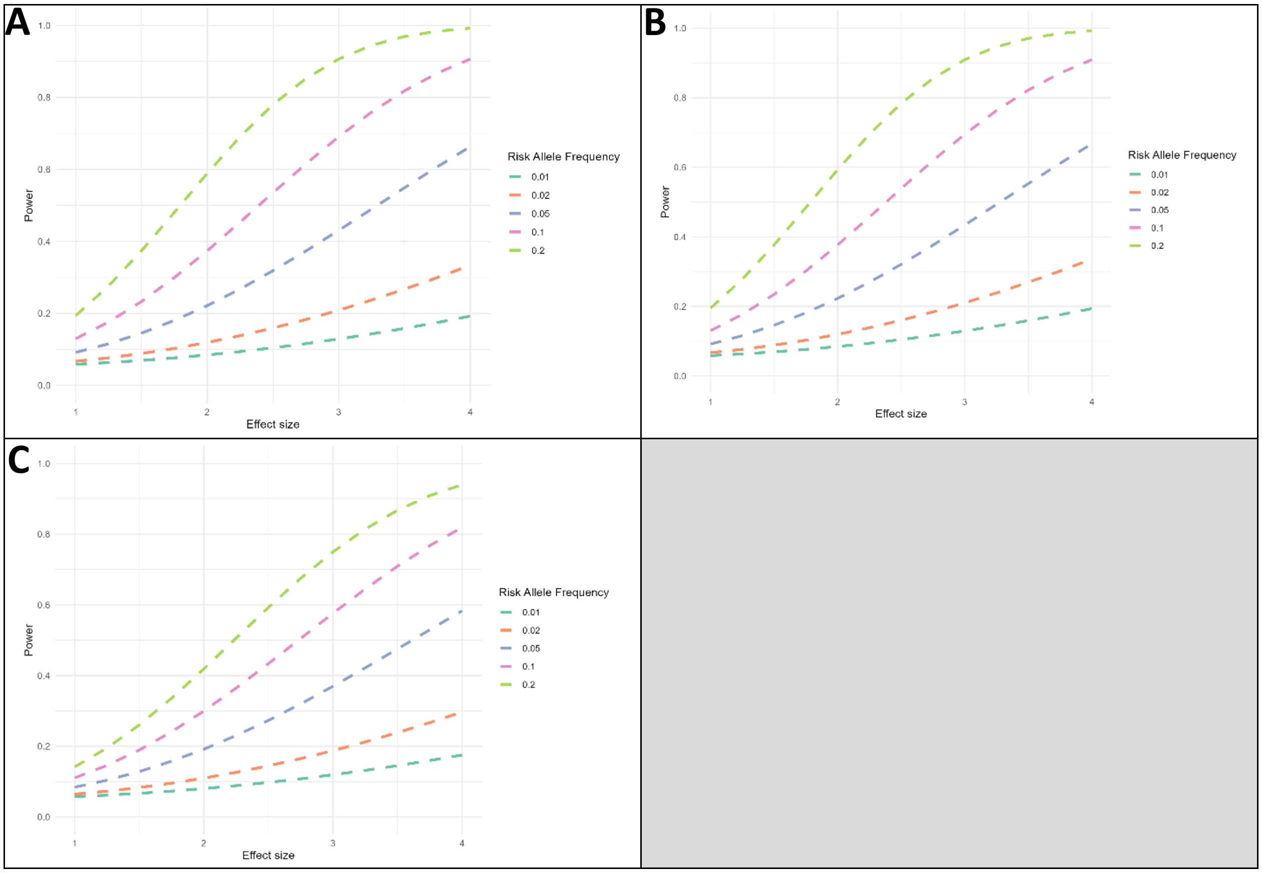

**Supplementary Figure S7 –** Statistical power analysis curves for the 1-year eGFR GWAS in recipients (A, additive model), donors (B, additive model) and pairs (C, dominant model) for the KiT-GENIE discovery cohort.

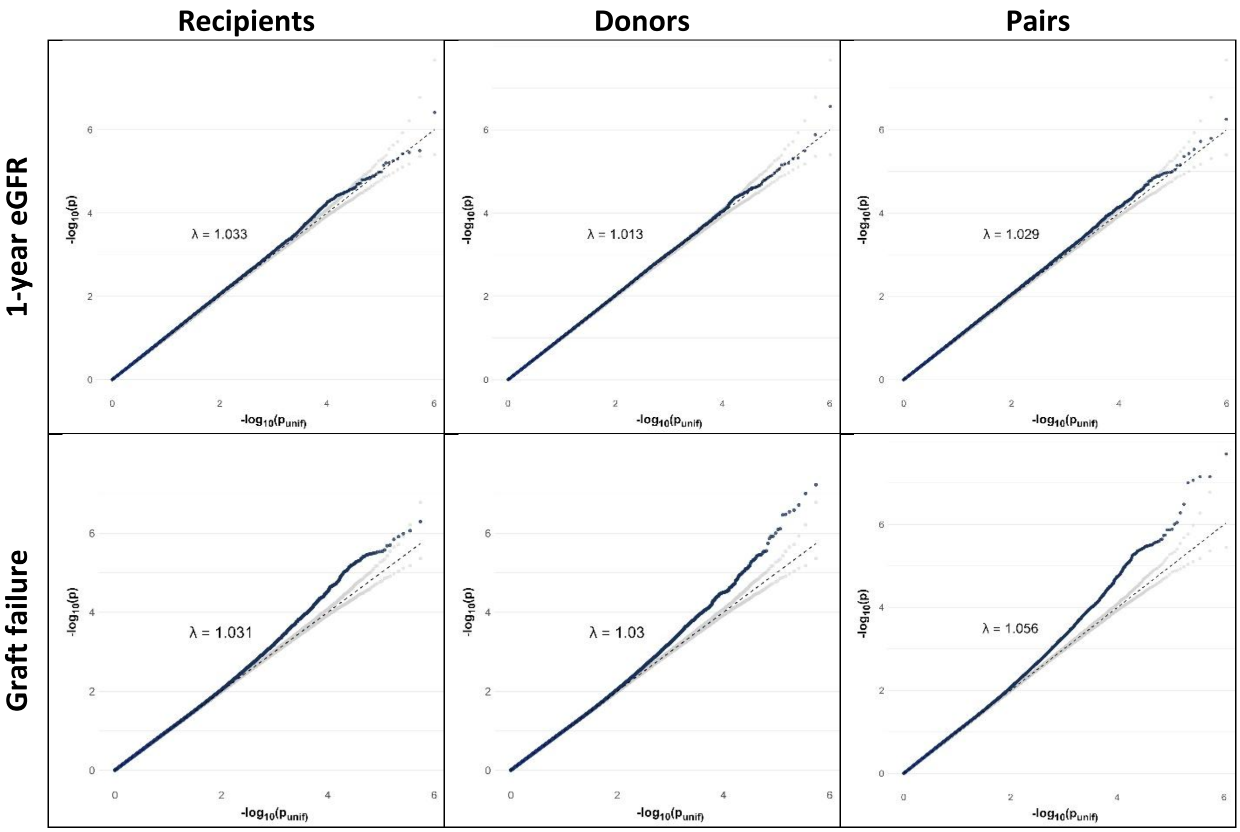

**Supplementary Figure S8 –** SNP association tests QQplots for the KiT-GENIE discovery cohort. Minor inflation was detected for graft loss while no inflation nor deflation was found for 1-year eGFR.

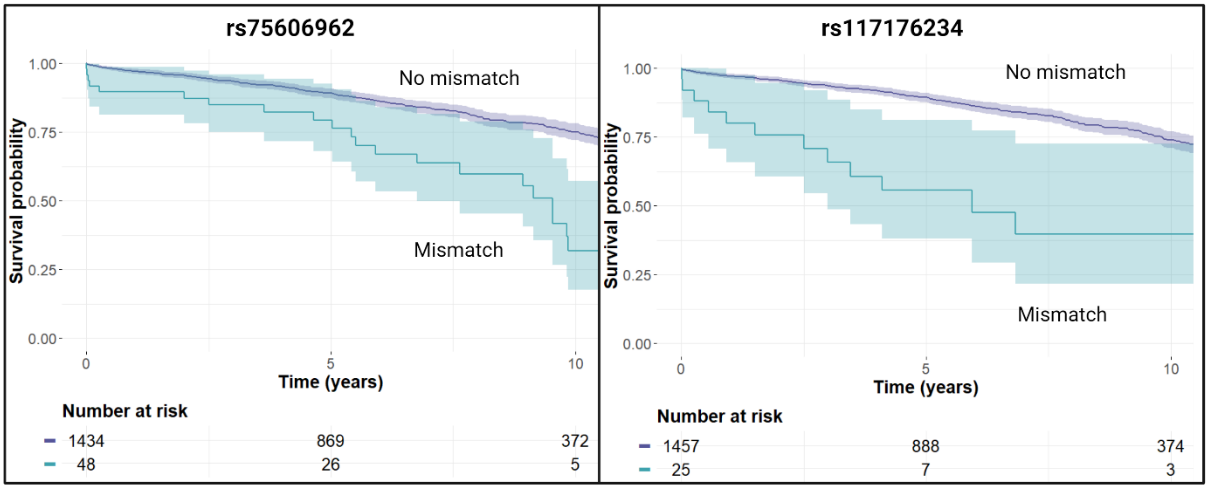

**Supplementary Figure S9 –** Kaplan Meier plots comparing kidney graft survival between matched and mismatched groups for the chromosome 10 (left panel) and chromosome 21 (right panel) top signals in the KiT-GENIE cohort. There is consistently a significantly worse survival for the transplants presenting a mismatch according to the log-rank test (p<0.0001).

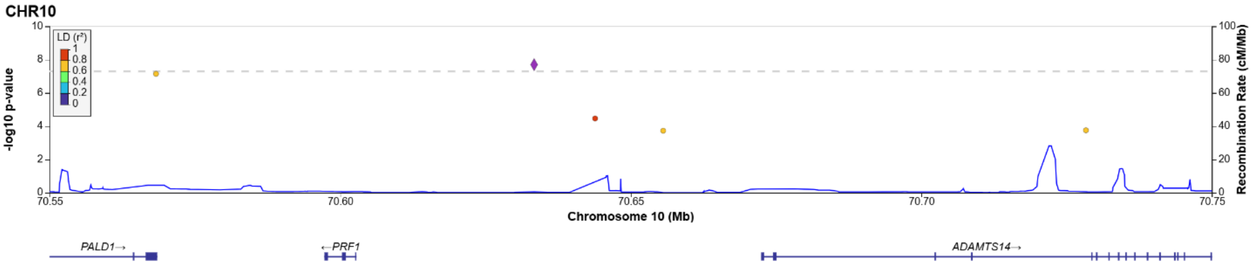

**Supplementary Figure S10 –** Locus Zoom plots for the unreplicated chr10 signal in the KiT-GENIE discovery cohort.

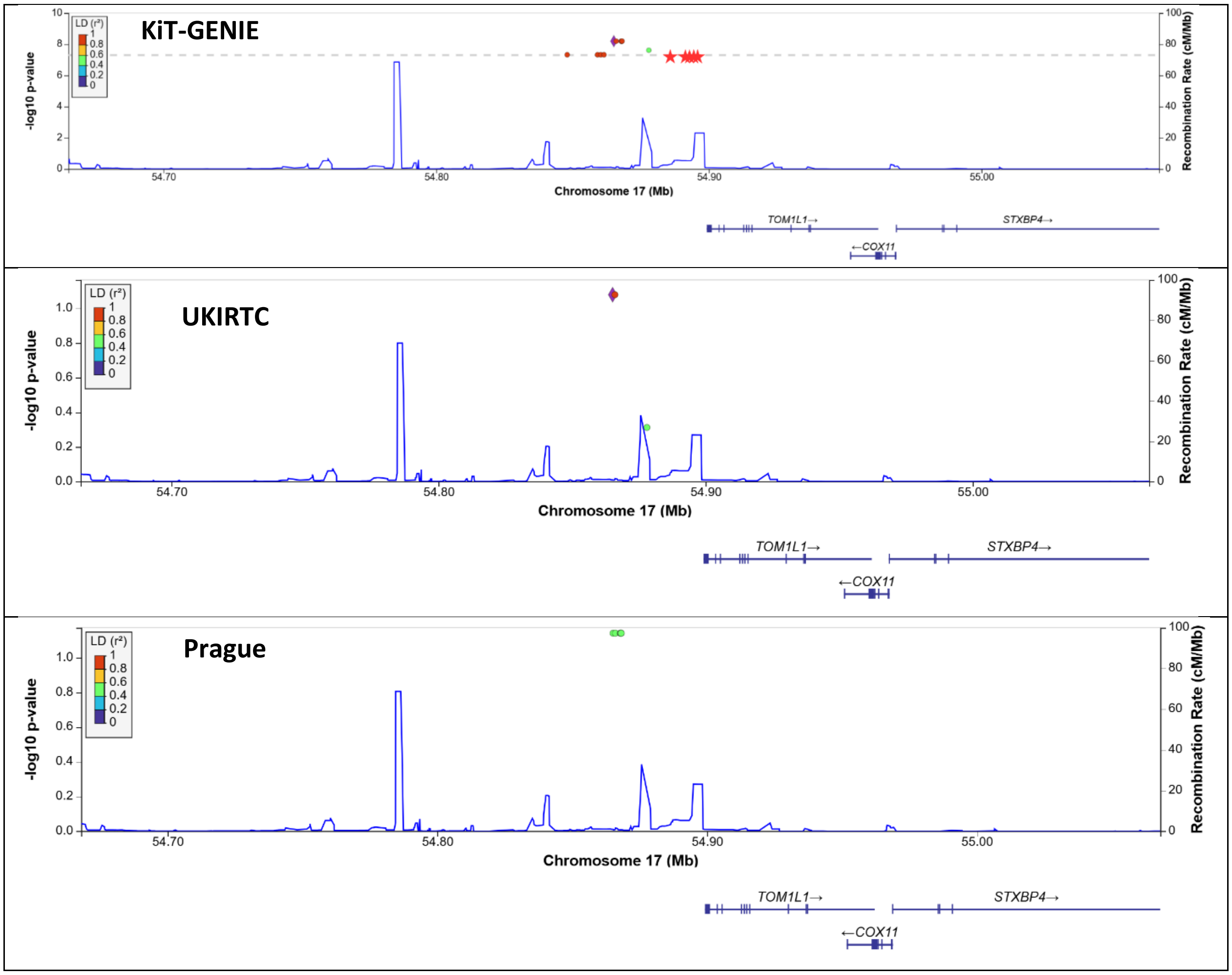

**Supplementary Figure S11 –** Locus Zoom plots for the replicated chr17 signal in the KiT-GENIE discovery and the validation cohorts. Red stars on the top row plot (KiT-GENIE) correspond to kidney tissue specific eQTLs (reported in Liu *et al* Nat Genet. 2022).

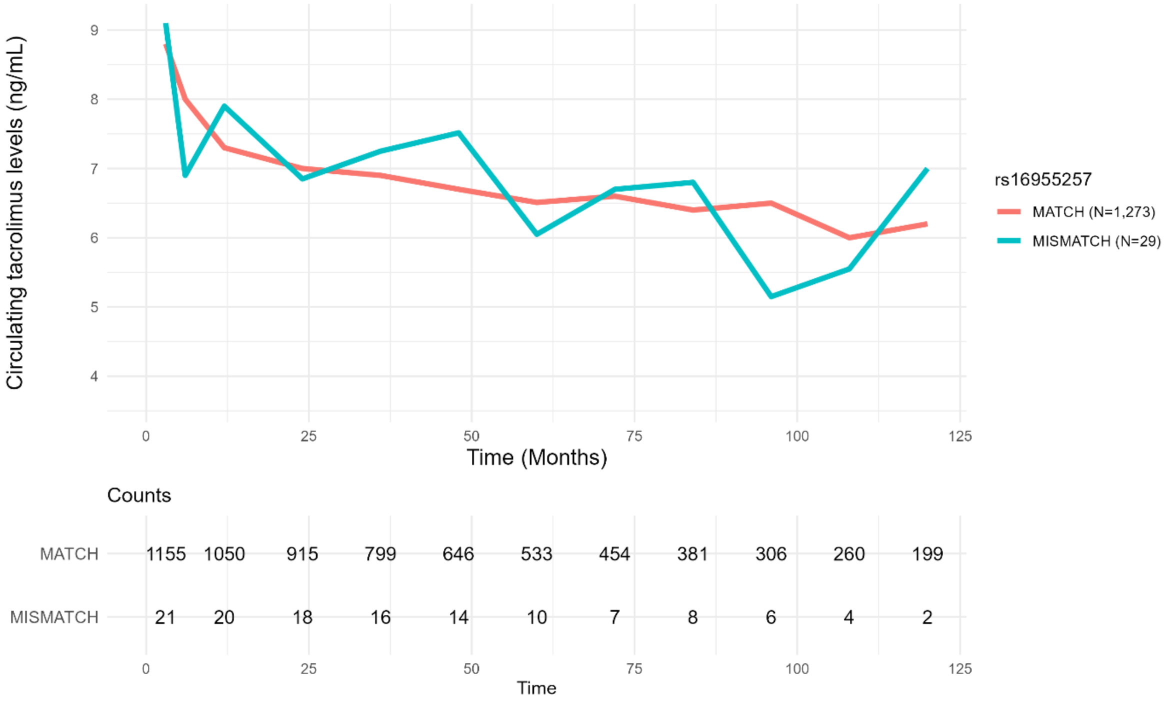

**Supplementary Figure S12 –** Evolution of the Tacrolimus circulating levels between the chr17 rs16955257 matched (red) and mismatched (blue) groups over the follow-up (missing data for N=180 patients out of 1,482). For each post-transplant visit (at 3, 6, 12 months and then annually thereafter), each curve represents the median Tacrolimus circulating levels within a group. Overall, median Tacrolimus circulating levels was of 7.1 ng/mL in KiT-GENIE and there was no major difference in the IS follow-up or adherence between the matched and mismatched groups. Data beyond 10 years is not shown here as the mismatch group sample size was very low.

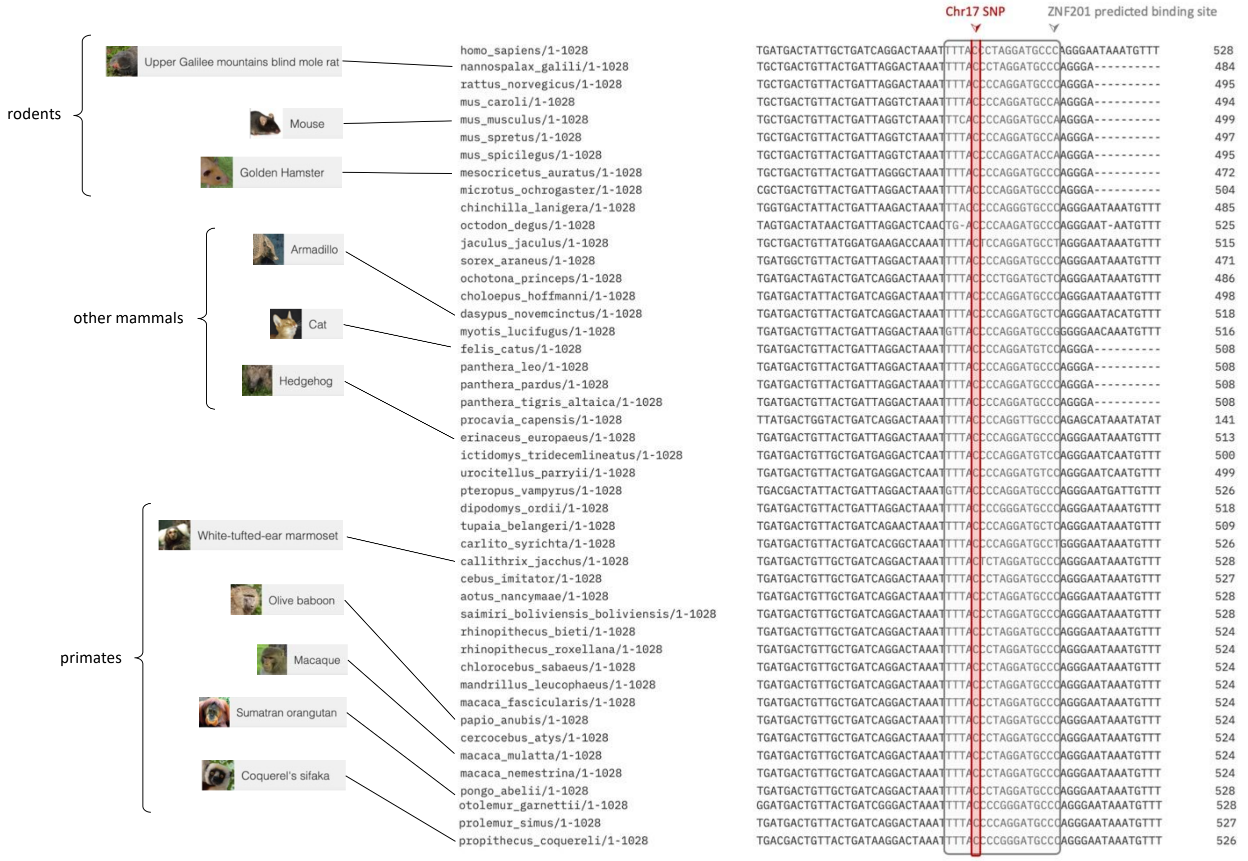

**Supplementary Figure S13** – Multiple sequence alignment for the rs16955257 (in red) flanking regions across 45 non-human species. Sequences were retrieved from the RefSeq database and alignement was performed with the Clustal-O algorithm.

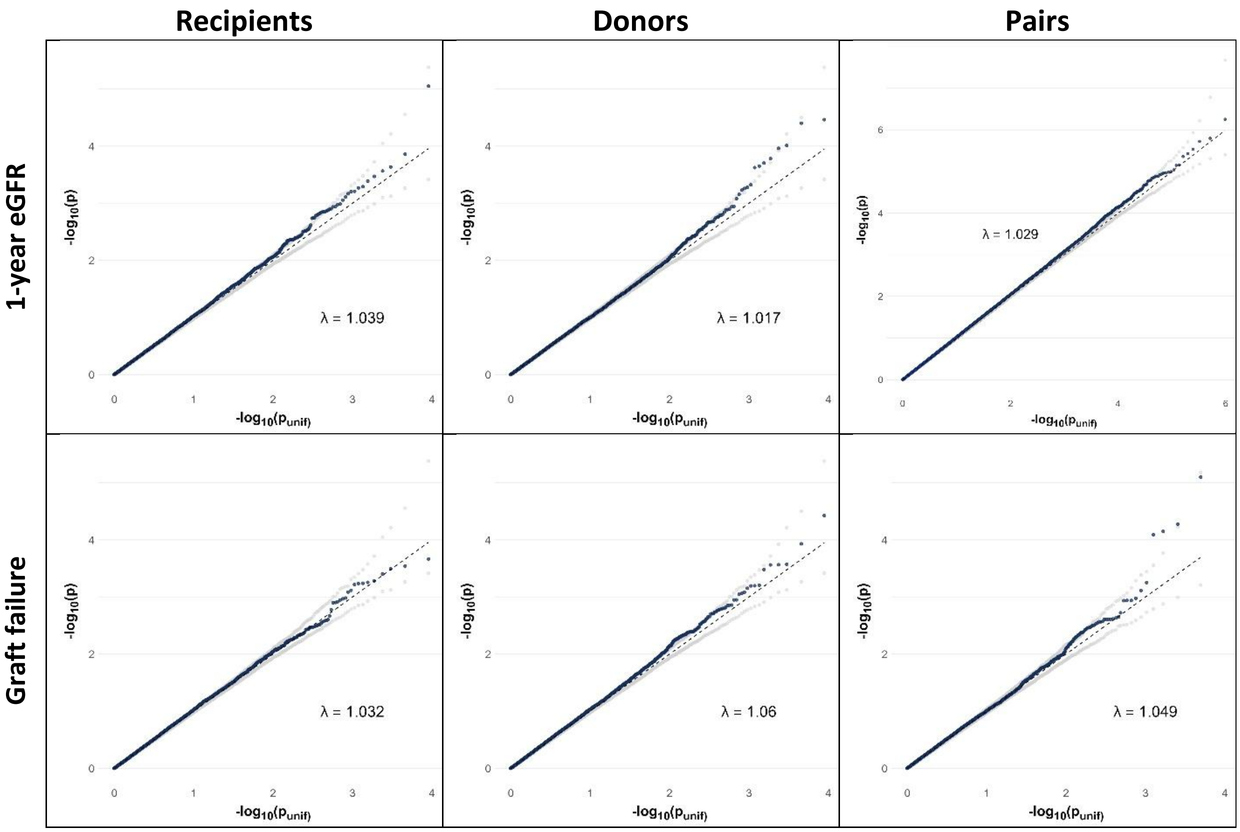

**Supplementary Figure S14 –** CNV association tests QQplots in the KiT-GENIE discovery cohort. No inflation nor deflation was detected.

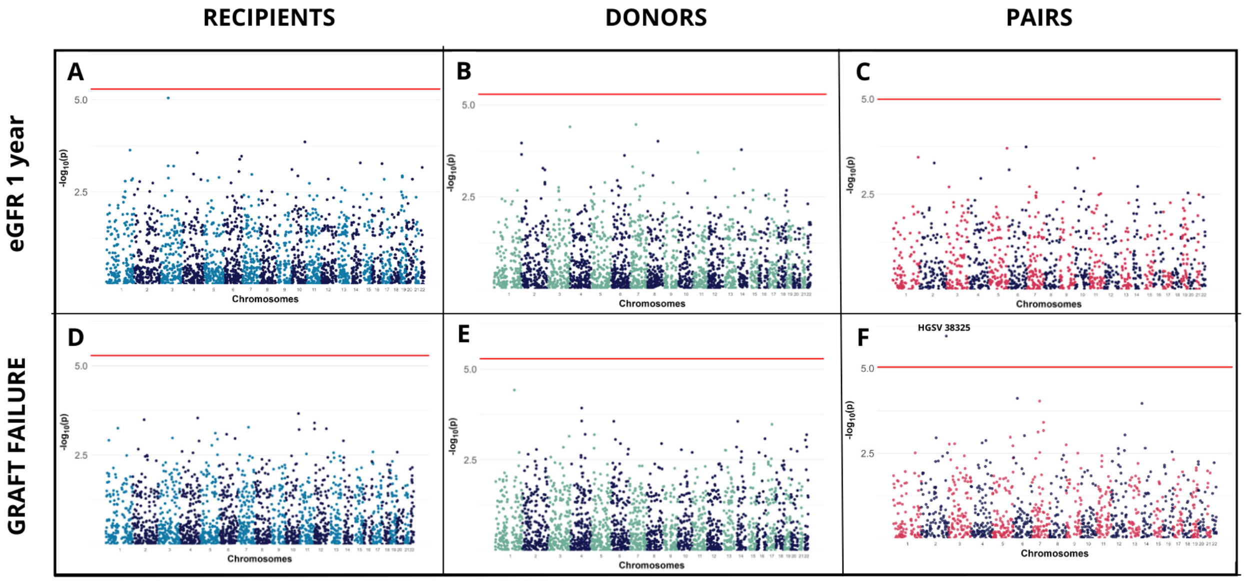

**Supplementary Figure S15** - Summary of CNV association tests - Results of the association tests between recipients' CNVs (panels A, D), donors' CNVs (panels B, E), and donor-recipient CNV mismatches (panels C, F) with 1-year eGFR (panels A-C), and time-to-graft failure (panels D-F) in the KiT-GENIE cohort. The x-axis represents the CNVs' position in the genome and the y-axis corresponds to the log-transformed p-values of the association tests. The Bonferroni genome-wide significance threshold is shown as a solid red line. Regression models were corrected for graft year, graft rank, recipient's and donor's age and sex, donor's type (living or deceased), *HLA* epitopic mismatches and PCA-computed genetic ancestry. A significant association was identified between a CNV mismatch in chromosome 2 and time-to-graft failure (panel F, $p=1.1x10^{-6}$, HR = 4.5 [2.5 – 8.4]).

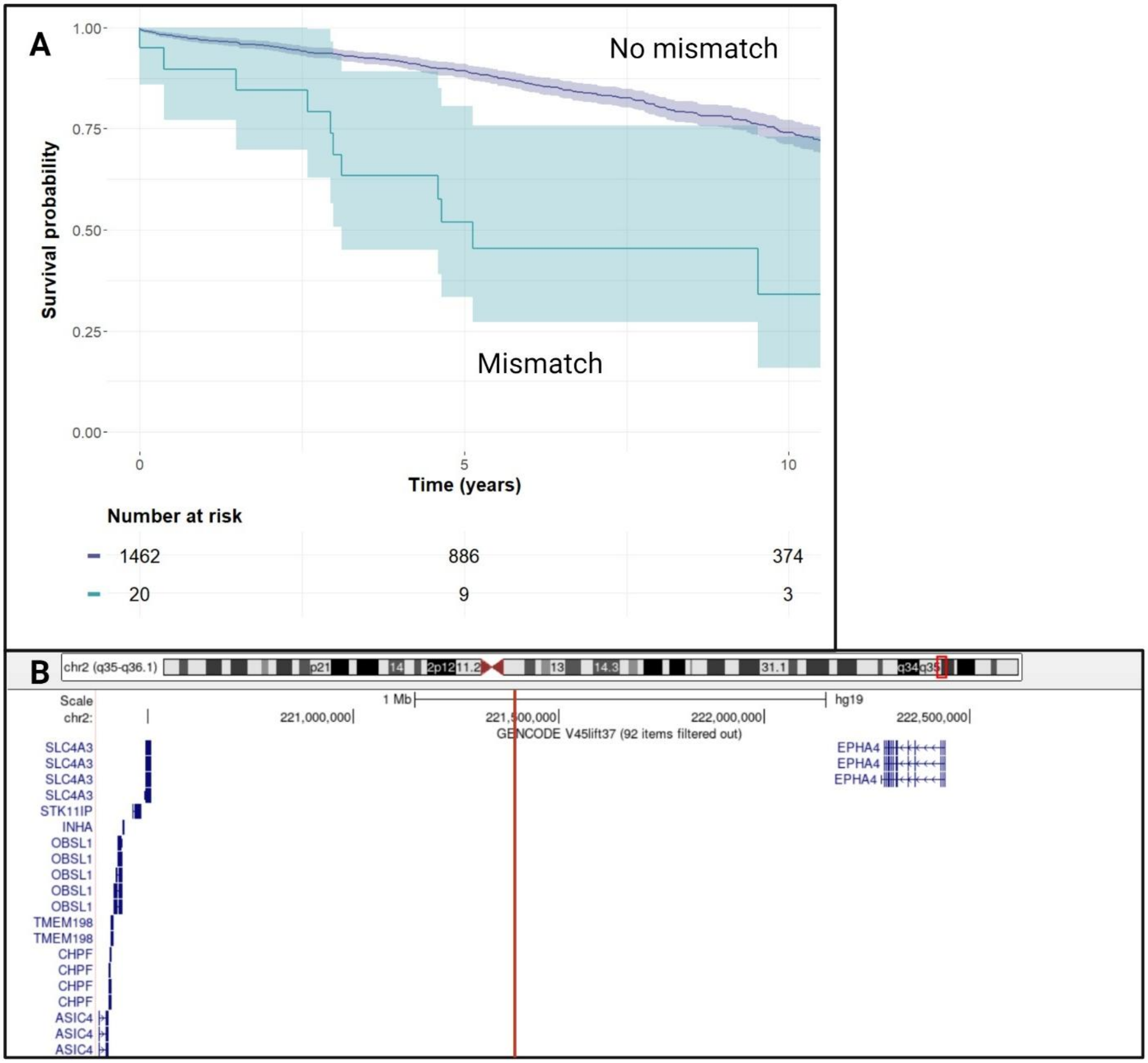

**Supplementary Figure S16 -** A chromosome 2 duplication mismatch (CNV tagged by the rs2392713 SNP) was significantly associated with time-to-kidney graft failure in the KiT-GENIE discovery cohort (p=1.1x10-6, HR=4.5 [2.5-8.4]). (A) The Kaplan Meier plot displays that mismatched pairs have a worse graft survival than matched pairs. (B) UCSC genome browser view of the genetic region surrounding the duplication (solid red line). The insertion lies in an intergenic region 900 kb downstream the EPHA4 gene.

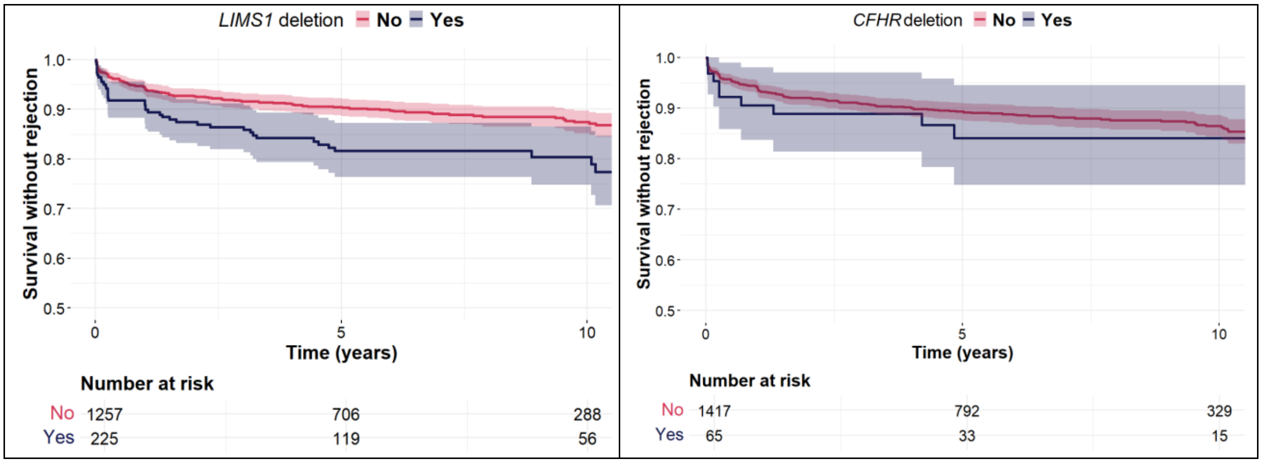

**Supplementary Figure S17 –** Survival curves stratified by the deletion-tagging SNP mismatches in the *LIMS1* locus (left panel) and the *CFHR* locus (right panel) for the KiT-GENIE cohort. The *LIMS1* mismatch confers a significant worse risk of rejection (p=0.001), while we did not find a significant effect of the *CFHR* locus mismatch on rejection (p=0.69).

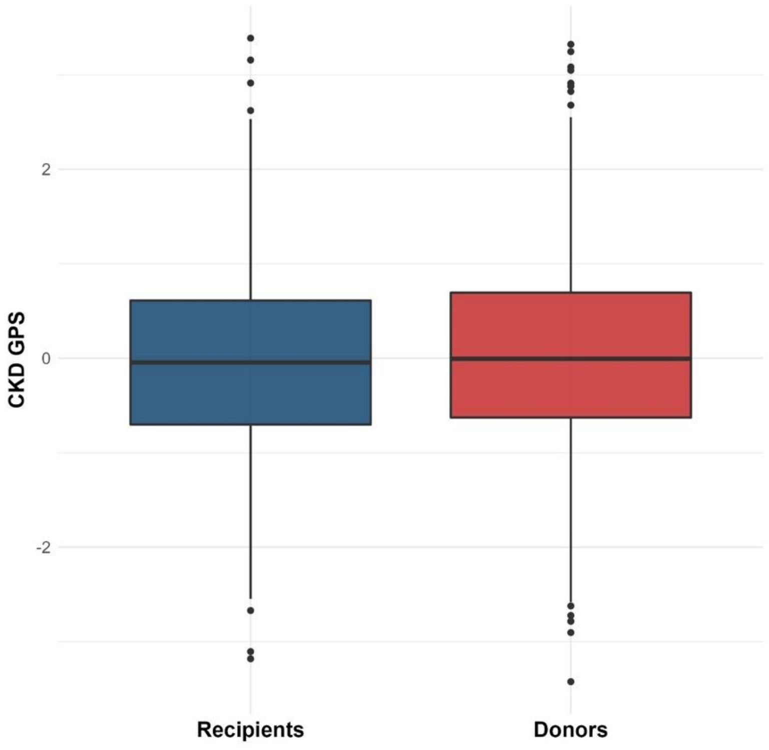

**Supplementary Figure S18 –** Boxplot comparing the distribution of the CKD GPS developed by Khan *et al*. between recipients (considered as CKD cases) and unrelated donors (considered as controls) in the KiT-GENIE cohort. We did not observe any major difference between recipients and donors (p=0.7).

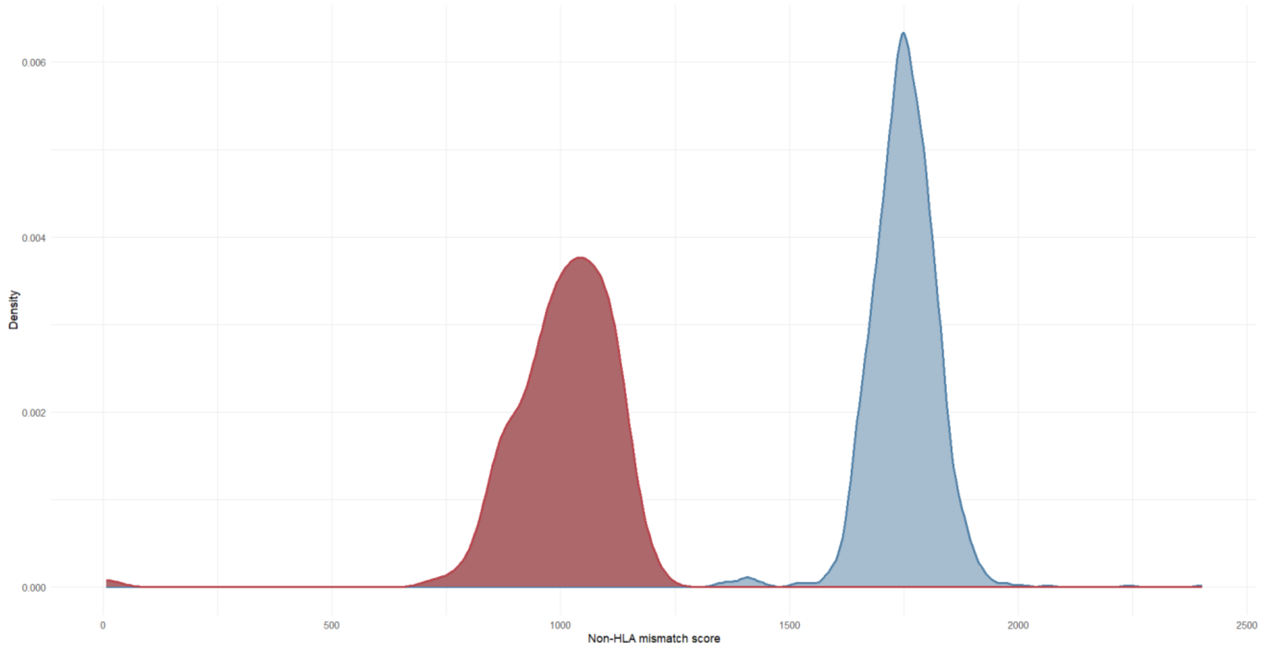

**Supplementary Figure S19** - Distribution of the non-HLA mismatch score (before normalization) in unrelated (blue, coefficient of variation = 4.3%) and related (red, coefficient of variation = 12.2%) donor-recipient pairs from the KiT-GENIE cohort. The mismatch score is significantly lower in related individuals, as expected (Pearson correlation coefficient of -0.98 with the donor-recipient identity-by-descent value).
